# Evidence of lesions from Epstein-Barr virus infection in human breast cancer genomes

**DOI:** 10.1101/2024.06.24.24309410

**Authors:** Bernard Friedenson

## Abstract

Epstein-Barr virus (EBV) infects essentially all humans and provides no benefit. EBV can cause nasopharyngeal cancer (NPC), Burkitt’s lymphoma (BL), and perhaps breast cancer. Breast tissues from patients with breast cancer are more likely to be EBV-positive than tissues from healthy controls. However, EBV is not a proven cause of breast cancer because the tissues are not consistently EBV-positive. If EBV causes breast cancer, it would have to do it without an active infection. Other cancers with known viral origins do not require continuing presence of the virus. However, the "hit and run" theory is difficult to test for breast cancer without a proven EBV connection.

Here, I test this theory with multiple independent bioinformatic analyses. First, hundreds of breast cancer genomes contained characteristic methylation scars that indicate a cleared EBV infection. The genomes had further differential hypermethylation near positions where EBV reprograms normal cells into malignancy. Second, genomes from EBV cancers and breast cancers inactivated the same tumor-suppressive mechanisms. Third, deletions were identified on chromosome 3p in EBV cancers that shift cells to oxidative glycolysis, a prominent breast cancer phenotype known as the Warburg effect. Similar 3p deletions were found in breast cancer genomes. Fourth, somatic hypermutation clusters in EBV-cancers marked genome positions in breast cancers near translocations and focal oncogene amplification. EBV deregulation of deaminase and estrogen-induced topoisomerase explain these translocation breakpoints. Fifth, several alternate explanations for these results were ruled out. Finally, only limited segments of EBV DNA matched the human genome, making it possible that a childhood vaccine would end breast cancer.

## Introduction

Saliva, blood, breast milk [1], and semen transmit Epstein-Barr virus (EBV). This transmission is so effective that virtually all humans acquire EBV infection at some point in their lives [2]. The infection has no known benefit to the host.

The virus creates reservoirs in the throat and lymph nodes, sometimes targeting the back of the throat, where it can cause nasopharyngeal cancer (NPC) and the lymphoid tissue, where it can cause Burkitt’s lymphoma (BL). NPC is the best studied example of an EBV cancer in epithelial tissue. Detection of EBV DNA in plasma samples is a nearly 100% accurate screen for NPC [3]. In lymphoid tissue, EBV transforms B-cells in culture [4] and antibodies against viral capsid antigen predict BL [5]. Both NPC and BL are therefore unequivocal examples of cancers caused by EBV. In both cancers, EBV attaches to human chromosomes as a circular episome and replicates along with host DNA. The EBV episome is exceedingly difficult to maintain in NPC cell culture because malignant cells lose the virus [6]. BL cell lines also lose or inactivate EBV episomes, but they are more stable than in NPC [6, 7]. The spontaneous loss of viral episomes shows that EBV can predispose to malignancy but is not required after transformation occurs.

Even though the virus can disappear, it still leaves evidence of infection behind. EBV-infected oral keratinocytes that expressed telomerase but were not malignant provide an example of this evidence. These cells have hypermethylated CpG islands as epigenetic scars. The methyl groups remain even if repeated host cell divisions cause the loss of the virus. These epigenetic alterations change gene expression, delay differentiation and can lead to an invasive phenotype [8, 9]. The mechanism for these changes included EBV infection upregulating DNA methyltransferases (DNMT3A, DNMT3B, and DNMT1). Non-malignant keratinocytes were more stable than carcinoma cells, but EBV infection reiterated methylation patterns found in NPC and gastric carcinoma [9]. Thus, patterns of hypermethylation provide a marker for past EBV infection, even if the infection disappears.

EBV reprogramming of the host epigenetic system causes massive changes in methylation characteristic of NPC [10, 11] NPC has hypermethylation in at least 80% of cases [12]. In the remaining 20% of NPC, subtle changes in specificity and activity of methyltransferases DNMT3A vs. DNMT3B may account for hypomethylation [13]. EBV also suppresses the removal of methyl groups by demethylation pathways TET-TDG-BER and APOBEC/AID-MBD4. These changes alter chromatin exposure [12] . Like NPC, BL causes epigenetic reprogramming by silencing at least one critical transcription factor [14]. BL and NPC both have inappropriate activation of the inflammatory NFKB pathway [15, 16], causing massive immune deregulation. Both these EBV cancers have significant off-target effects of activation induced deaminase (AID) which causes local somatic hypermutation (SHM) [15]. BL includes gene rearrangements which often deregulate the MYC gene. Most viral genes disappear from BL, perhaps by hijacking and perverting functions from host genes [17]. Both NPC and BL have forms in which EBV is undetectable, compatible with loss of virus as the cancer progresses. Thus, both NPC and BL provide data that support one current model for how EBV causes cancer. The model suggests that inactivation of tumor suppressors and immune defenses [16] occurs in premalignant lesions. These deficits then allow latent EBV in reservoirs to trigger critical epigenetic changes throughout the lesions. EBV reprograms the human regulatory epigenome, changes chromatin accessibility, then cellular differentiation and proliferation [12]. The continuing presence of the virus is not required once cells become malignant.

In contrast to its role in NPC and BL, a role for EBV in breast cancer is not broadly accepted [18]. Epidemiologic studies have shown that evidence of current EBV infection is about five-fold more likely in breast cancer tissues than in non-malignant controls [19]. However, only 4 of 30 studies convincingly demonstrated the presence of EBV while 14 studies convincingly demonstrated its absence [20]. The inconsistent detection of EBV in breast cancers leads to suggestions that EBV predisposes mammary epithelial cells to breast cancer, but the virus is not required after the cells become malignant [21]. Breast cancer initiation is more likely in individuals when near-universal EBV infection escapes from control such as by immunocompromise, but because replicating cells can clear EBV, active EBV infection cannot always be detected. Cancer in the absence of its origins describes several other viral or microbial cancers [22, 23]. However, there is currently no way to determine whether this hit and run mechanism occurs in breast cancer.

The aim of the present study was to determine whether oncogenic lesions left by EBV infection tie a lost EBV infection to breast cancer. The experiments were multi-pronged bioinformatic comparisons of breast cancer genomes to EBV infected cells and model EBV cancers. Characteristic epigenetic, metabolic, and chromosome changes that EBV leaves behind in infected host cells converge on the idea that EBV mediated damage is a significant driver for human breast cancer. Five independent lines of evidence support the conclusion that EBV predisposes breast tissue to malignancy but is not required once transformation occurs. Only short EBV-like DNA sequences exist in the human genome and only match a small area of the EBV genome. This limited homology makes it possible that a childhood vaccine against products expressed by other segments of EBV DNA would effectively end breast cancer as we know it.

## Results

### Oral keratinocytes previously infected with EBV and breast cancer cells exhibit similar differential DNA methylation profiles

The aim of this experiment was to determine whether prior EBV infection may have left remnants of hypermethylation on breast cancer genomes. EBV-infected oral keratinocytes have abnormal methylation that remains even when the infection clears [9]. I compared the distributions of retained hypermethylation in these formerly infected cells to epigenetic marks in breast cancer cells [24] that are likely breast cancer drivers. The epigenetic marks in once-infected keratinocyte chromosomes globally predict which chromosomes are involved in breast cancer cells (**Fig. 1A**). The frequencies of which chromosomes have these epigenetic drivers correlate according to linear regression analysis (r=0.93,r2=0.87, p<0.0001), rejecting the null hypothesis that there is no relationship between chromosomes in EBV cancers and breast cancers. Two-or one sided Kolmogorov-Smirnov tests gave high p values (0.89 or 0.5 to 0.44), suggesting any differences in the two distributions are likely due to chance. Similarly, Spearman’s rank correlation gave a correlation coefficient of 0.88 (CI=0.72-0.95). Although these results are not a detailed comparison of methylation sites, they were the first hints of a potential relationship between EBV epigenetic reprogramming and breast cancer drivers.

**Fig. 1A.**
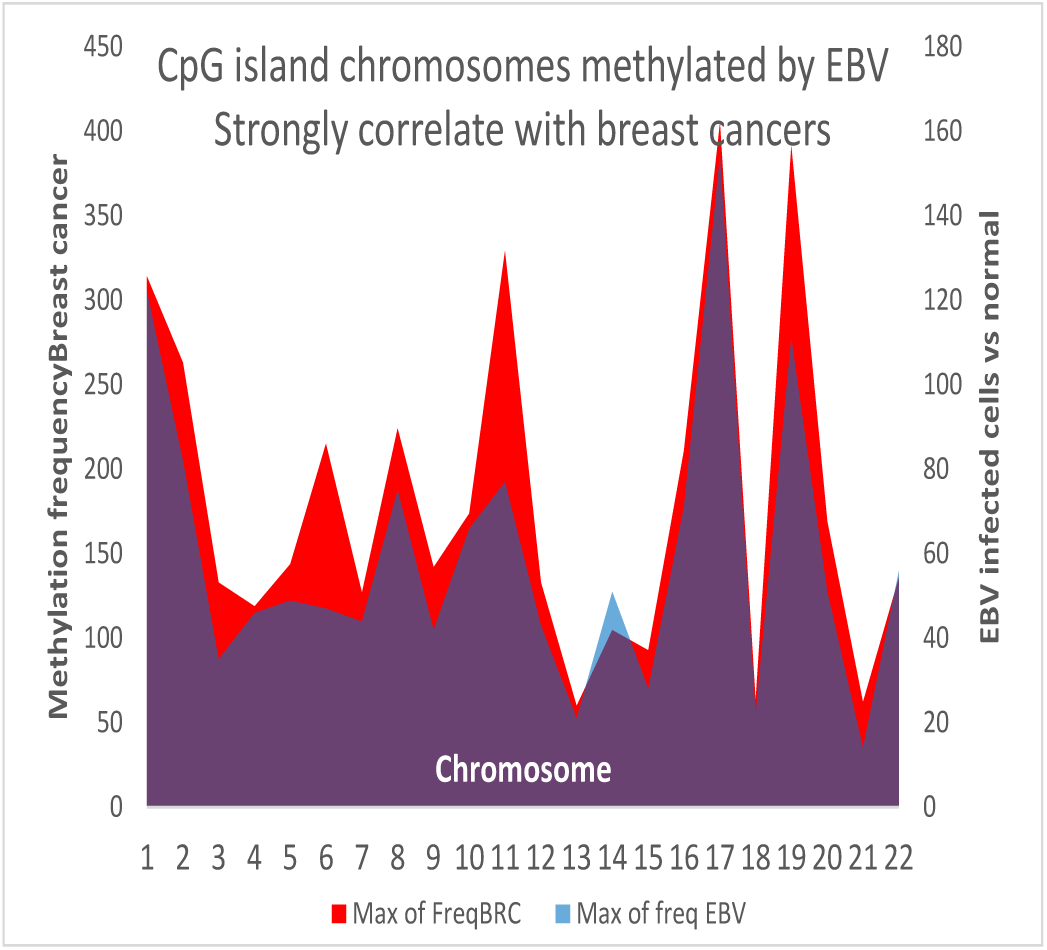
The frequency of differential methylation of chromosomes in oral keratinocytes that have cleared EBV correlates with the frequency of differential methylation in breast cancer chromosomes.

To examine the relationship between breast cancer drivers and EBV infected oral keratinocytes in greater detail, I compared the exact genome coordinates of abnormal methylation in both systems. I found breast cancers have at least eight chromosomes with methylation marks in about the same position as the formerly infected keratinocytes (**Fig. 1B**). Initially, the screening window was 200,000 bp. Then, the exact distances were calculated more closely for the ten genes methylated in breast cancer that were nearest to sites of abnormal methylation in oral keratinocytes that had lost their EBV infection (**Table 1**). Functional descriptions of the ten genes link them all to breast cancer development and metastasis (**Table 1**, last column, and supplementary references to **Table 1**). This result shows EBV infection has left characteristic marks on breast cancer cells, and these epigenetic scars can predispose to cancer.

**Fig. 1B.**
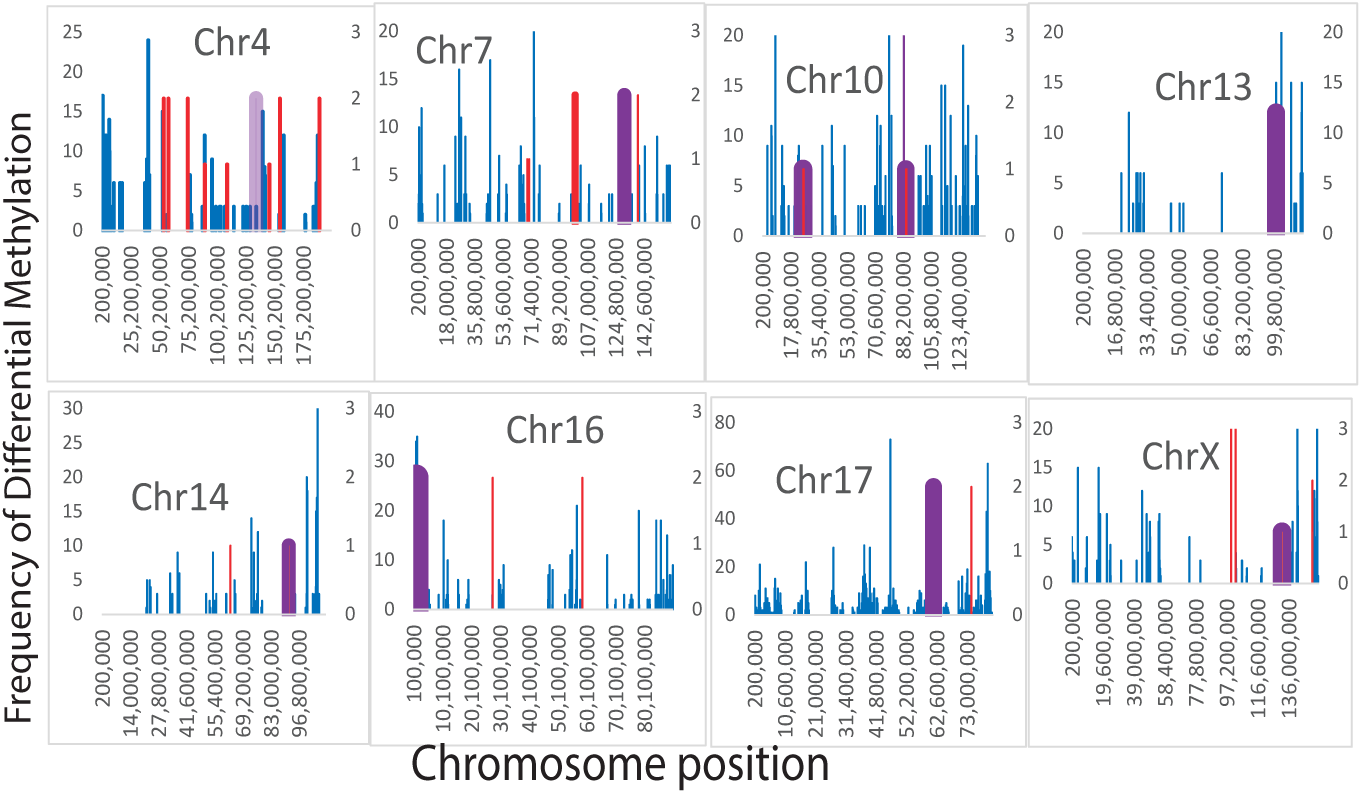
Differential methylations that persist after EBV infection clears from oral keratinocytes also occur in breast cancers. The frequencies of differential methylation in breast cancers cells (blue) and keratinocytes that have lost their EBV infection were compared. Chromosome positions that overlap are indicated in purple.

**Table 1.** Methylation sites that predispose to breast cancer are near aberrant methylation sites in infected keratinocytes that have lost their EBV infection.

| Gene showing persistent methylation after the clearance of EBV | Gene name | Chromosome coordinates (Hg19) | Distance (bp) from sites of hyper-methylation in breast cancers | Function, relation to breast cancer |
| --- | --- | --- | --- | --- |
| PCDH10 | Protocadherin-10 | 4: 134,070,449-134,129,761 | 945 | Cell-cell adhesion [87]. Inhibits cancer cell motility and cell migration. PCDH10 methylation is common in primary breast cancer and associated with poor survival [88]. |
| MEST | Mesoderm-specific transcript | 7: 130,126,016-130,146,306 | 4360 | Loss of imprinting has been linked to promoter switch and breast cancer metastasis [89] |
| GPR158 | Signal transduction, G-protein-coupled receptor | 10:25,463,930-25,891,158 | 465 | Estrogen withdrawal or antagonist exposure down-regulates GPR158 expression in estrogen-sensitive breast cancer. GPR158 may predict prognosis in breast and ovarian cancer [90] |
| RNLS | Renalase, FAD-dependent amine oxidase | 10:89,931,280-90,343,075 | 137 | Promotes survival with anti-inflammatory effects. Overexpressed in breast cancer [91] |
| ZIC2 | Zinc finger protein AIC family member 2 | 13:100,632,108 - 100,632,305; 100,632,306-100,632,466;<br>100,632,478-100,632,573;<br>100,633,028-100,633,249 | 789 | Regulates chromatin and transcription. Embryonic cell differentiation [92]<br>Downregulated in breast cancer [93] |
| CCDC88C | coiled-coil domain containing 88C | 14:91,737,667-91,884,164 | 4658 | Highly mutated in breast cancer. A tumor suppressor that is related to Wnt signaling [94] |
| FBXL16 | F-box and leucine-rich repeat protein 16 | 16:742,500-755,801 | 53 | Suppression is associated with aggressive, lethal breast cancers [95] |
| IL21R | interleukin 21 receptor | 16:27,413,495-27,463,363 | 8168 | Activates NK cells. Part of an immune prognostic gene panel that predicts breast cancer survival [96]. |
| CYB561 | cytochrome b561 | 17:61,509,665-61,524,000 | 8543 | Increases breast cancer cell proliferation, especially HER2+ breast cancer [97]. |
| HS6ST2 | heparan sulfate 6-O-sulfotransferase | X:131,760,043-132,095,398 | 199 | HS6ST2 encodes a heparan sulfate (HS) sulfotransferase gene. The encoded protein catalyzes the transfer of sulfate to HS. HS6ST2 regulates cell adhesion, |
|  | 2 |  |  | migration, growth, and differentiation. Low expression is related to poor breast cancer survival [98] |

### Sites of methylation in the EBV-related cancer NPC cluster around the sites of driver gene methylation in breast cancer

The next step was to determine whether the relationship between EBV infected cells and breast cancer extends to a relationship with EBV-mediated cancers. To test breast cancer for these EBV-type changes, I compared the positions of hypermethylation in EBV-positive NPC [12] to positions of hypermethylated driver mutations [24] in breast cancers. NPC hypermethylation positions agree with the positions of hypermethylation in breast cancer within or near genes that are being regulated for thirteen chromosomes (**Fig. 2A**). Not only did the positions of many NPC and breast cancer hypermethylated groups match closely, but it was possible that most chromosomes had the same distribution of methyl groups in chromosomes in these two different cancers. The frequency distributions of chromosome positions with abnormal methylated groups on NPC’s and breast cancers overlap at many loci on example chromosomes (**Fig. 2B**). These example chromosomes were selected because Mann-Whitney results (P>0.05) could not rule out identical distributions of differential hypermethylation.

**Fig 2A.**
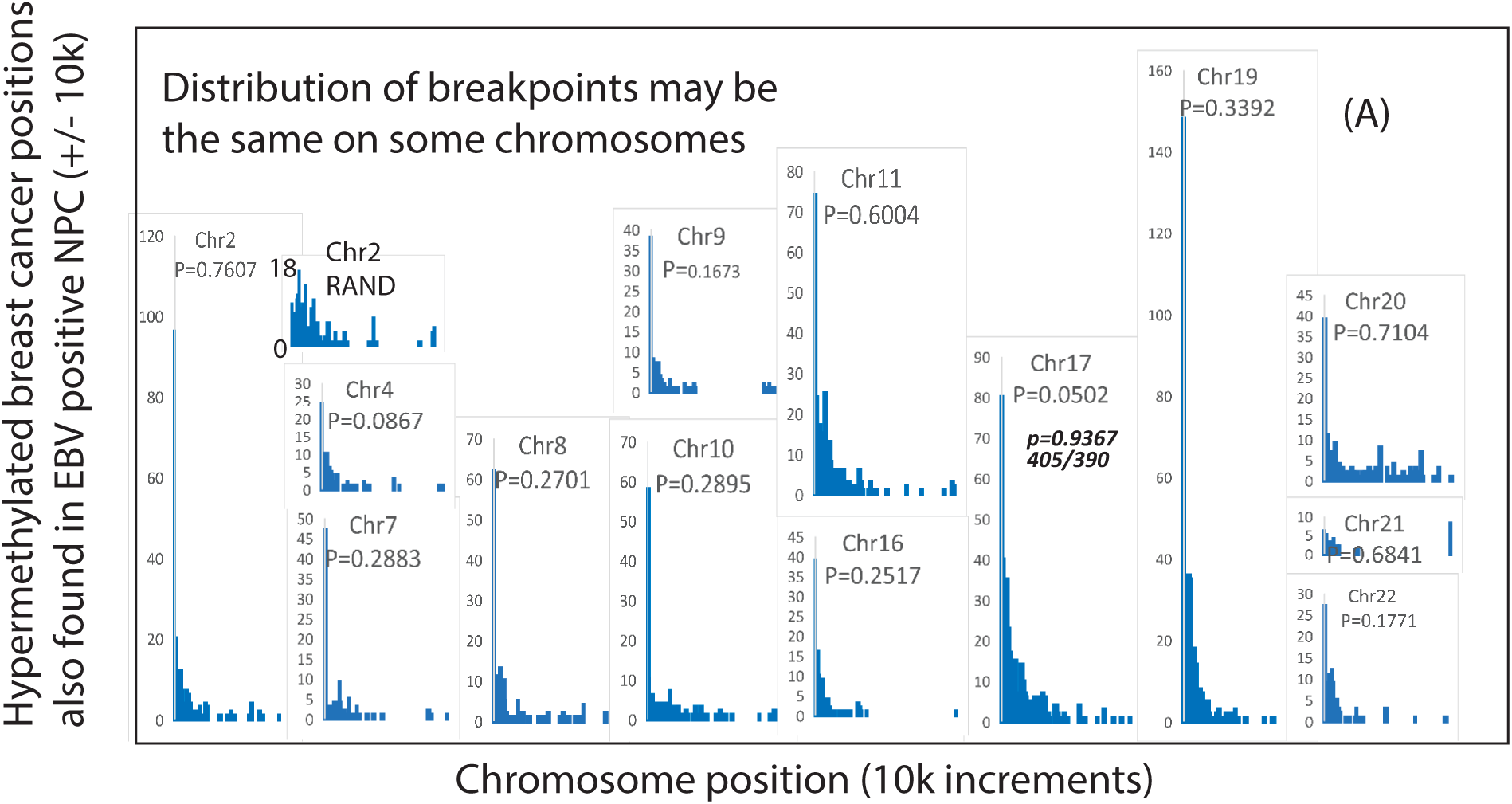
Positions of hypermethylation in NPC, an EBV-mediated cancer, relate to cis-hypermethylated positions in breast cancers. P values do not exclude the possibility that the hypermethylation positions in breast cancer and NPC are the same. Comparison to a normal random distribution with the same mean and standard deviation as the NPC data for chr2 is to the right of the chr2 graph and labeled “chr2RAND”.

**Fig 2B.**
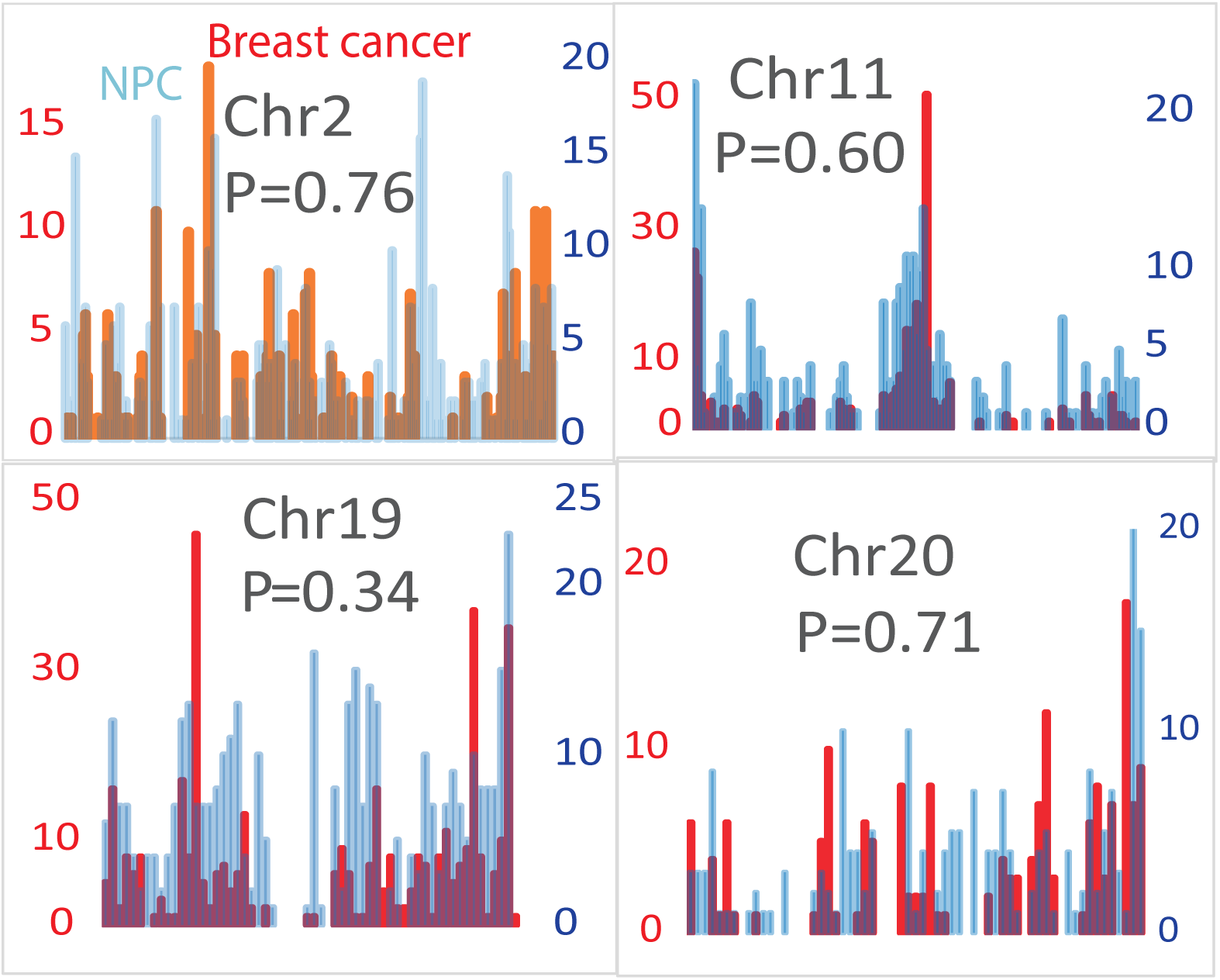
The frequency distributions of methyl groups overlap in NPC and breast cancer when the chromosomes are divided into bins of 1 million bp.

In cases where the distributions do not agree (P<0.05, **Fig 2C** ), the distributions of hypermethylation are the same in chromosome areas where matches occur (P values approach 1.0 in **Fig. 2C**). Both cancer sites involve a significant upregulation of DNMT3A, which catalyzes changes in the epigenome. To verify that the 10,000 base pairs window was not too large, breast cancer methylation sites were run against a random distribution with the same mean and standard deviation as the NPC data. Instead of nearly 100 positions that agreed, trials against two different random distributions gave values of 1 and 11 (**Fig 2A**). This result means that the agreement between NPC and breast cancer methylation positions is unlikely to occur purely by chance. These results show that massive methylation changes caused by EBV in NPC also occur in breast cancer.

**Fig. 2C.**
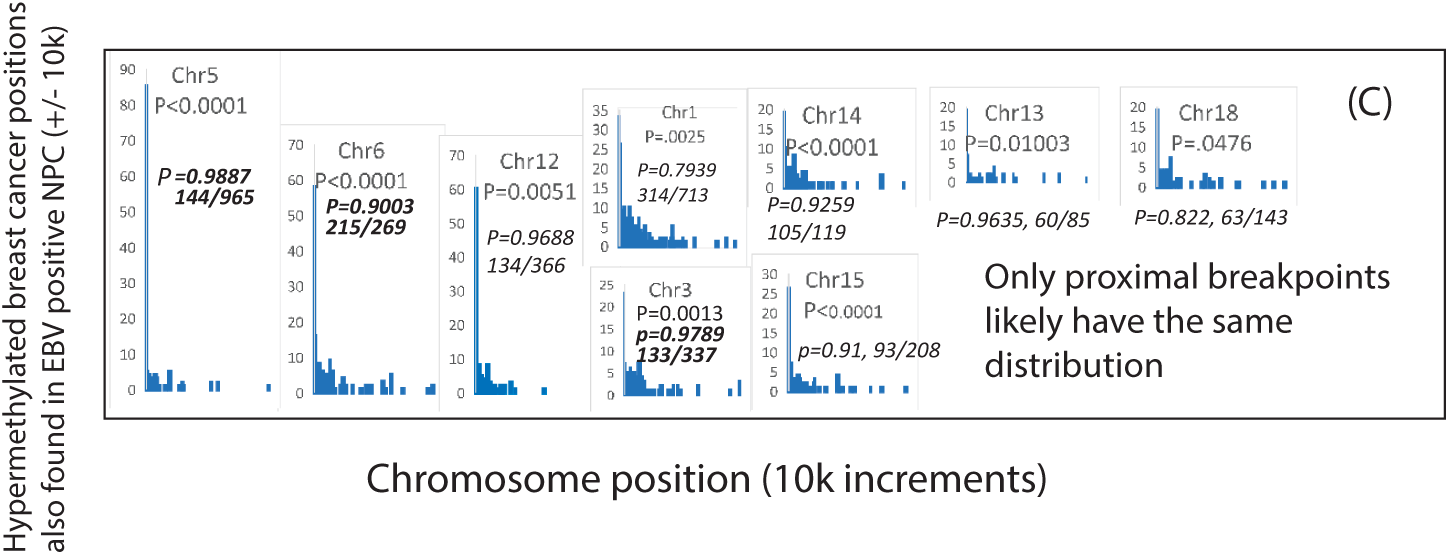
If P values suggested that the distributions of methylation positions in breast cancer and NPC were unrelated (P<0.05), probabilities were calculated that methylation positions within 10,000 base pairs of each other (fractions in bold type) came from the same distributions.

To determine if this agreement is more general, I tested BL, another cancer driven by EBV infection. Differential methylation positions near breast cancer drivers were the largest peak on all chromosomes. Again Mann-Whitney testing could not rule out the same hypermethylation distribution in BL and in breast cancer (P>0.05, **Fig. 3**). This result shows that many positions of differential methylation in a second EBV cancer are similar to those in breast cancers. These results support the possibility that EBV infection has driven the epigenetic changes in breast cancer.

**Fig. 3.**
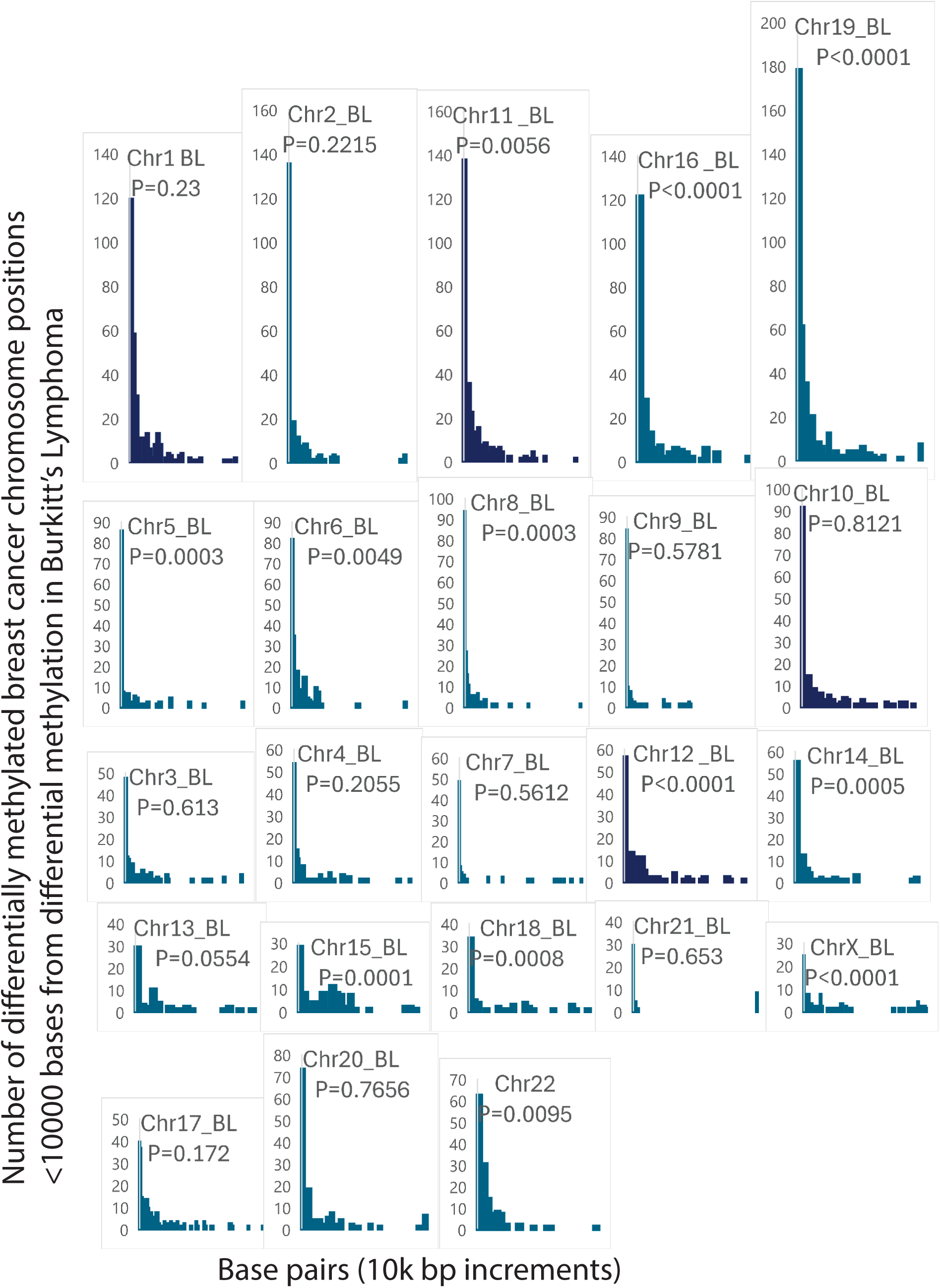
Differential methylation occurs on every chromosome at approximately the same positions in breast cancer and BL. Positions of differential methylation on every chromosome in 10k base pair increments (breast cancer vs. normal compared to BL vs. normal). P values are probabilities that the values come from the same distribution.

### Genes that control stem cell properties and antiviral immunity are key targets of hypermethylation that breast cancers share with EBV-driven cancers

To test whether the methylation changes had a plausible role in causing breast and EBV cancers, the functions of hypermethylated genes were researched. Hypermethylated genes in NPC were found to cluster around homeobox (HOX) on chromosome 17, which are also hypermethylated breast cancer driver genes (**Fig. 4**). These genes affect oncogenesis because they regulate stem cell differentiation and self-renewal [25]. Data from chromosome 18 (**Fig. 4**, bottom) show additional effects on chromatin, invasion, and hint at estrogen interaction (**Table 2**). These results provide examples of mechanisms that explain how differential methylation caused by EBV infection can predispose to breast cancer.

**Fig.4.**
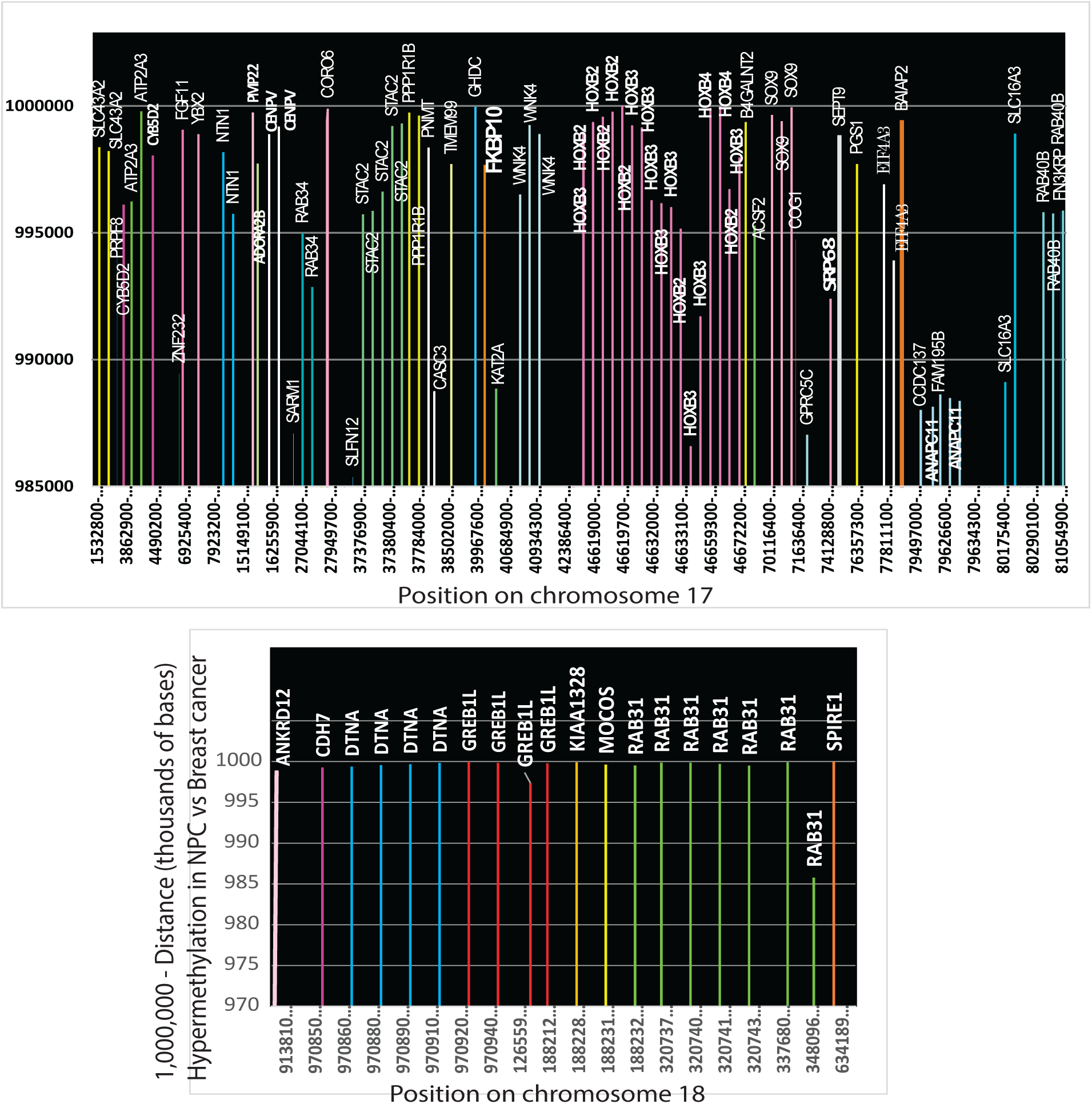
Genes hypermethylated in NPC cluster around cis-regulated genes hypermethylated in breast cancer. Data from chromosome 17 and chromosome 18 are shown as examples.

**Table 2.**
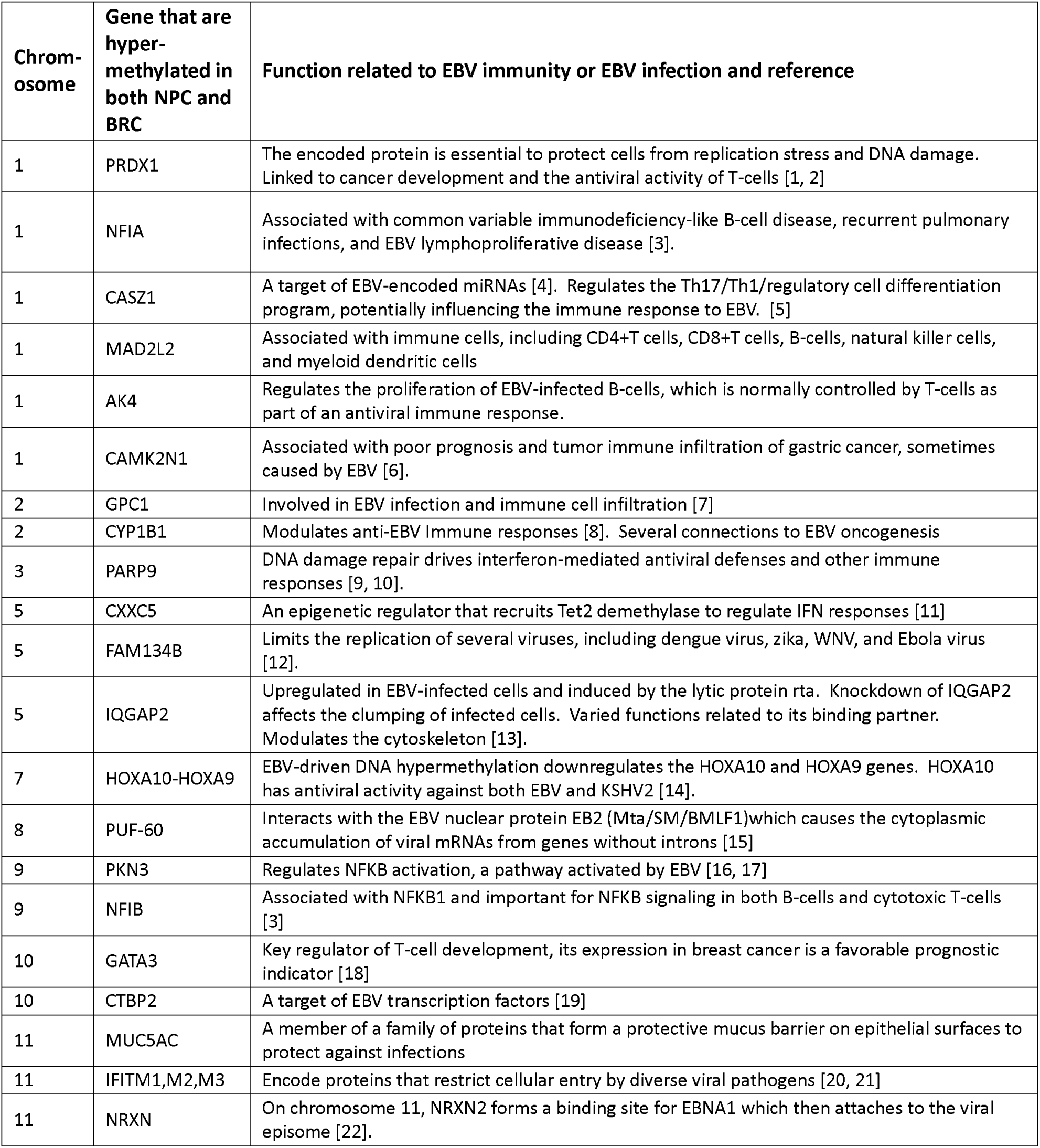
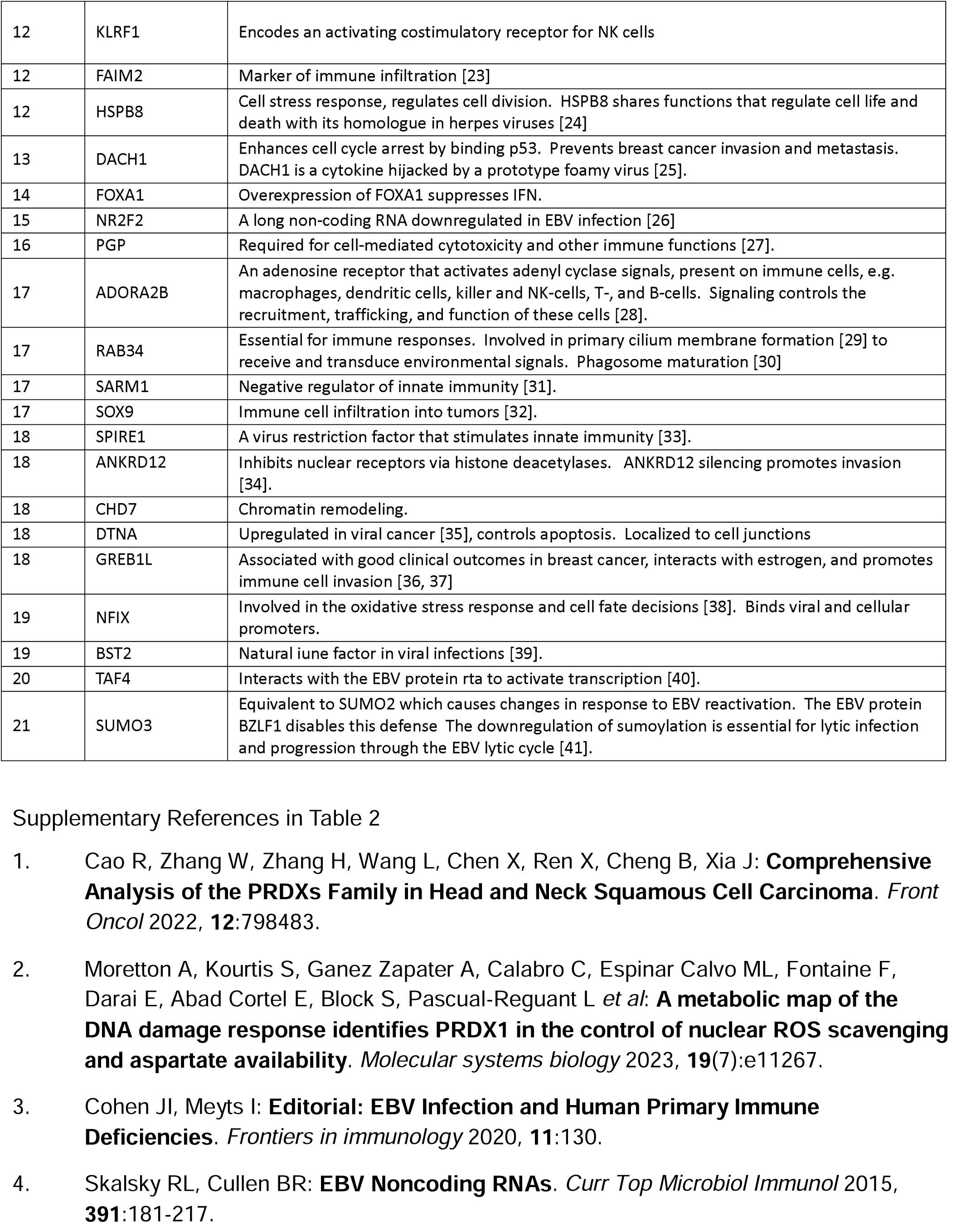
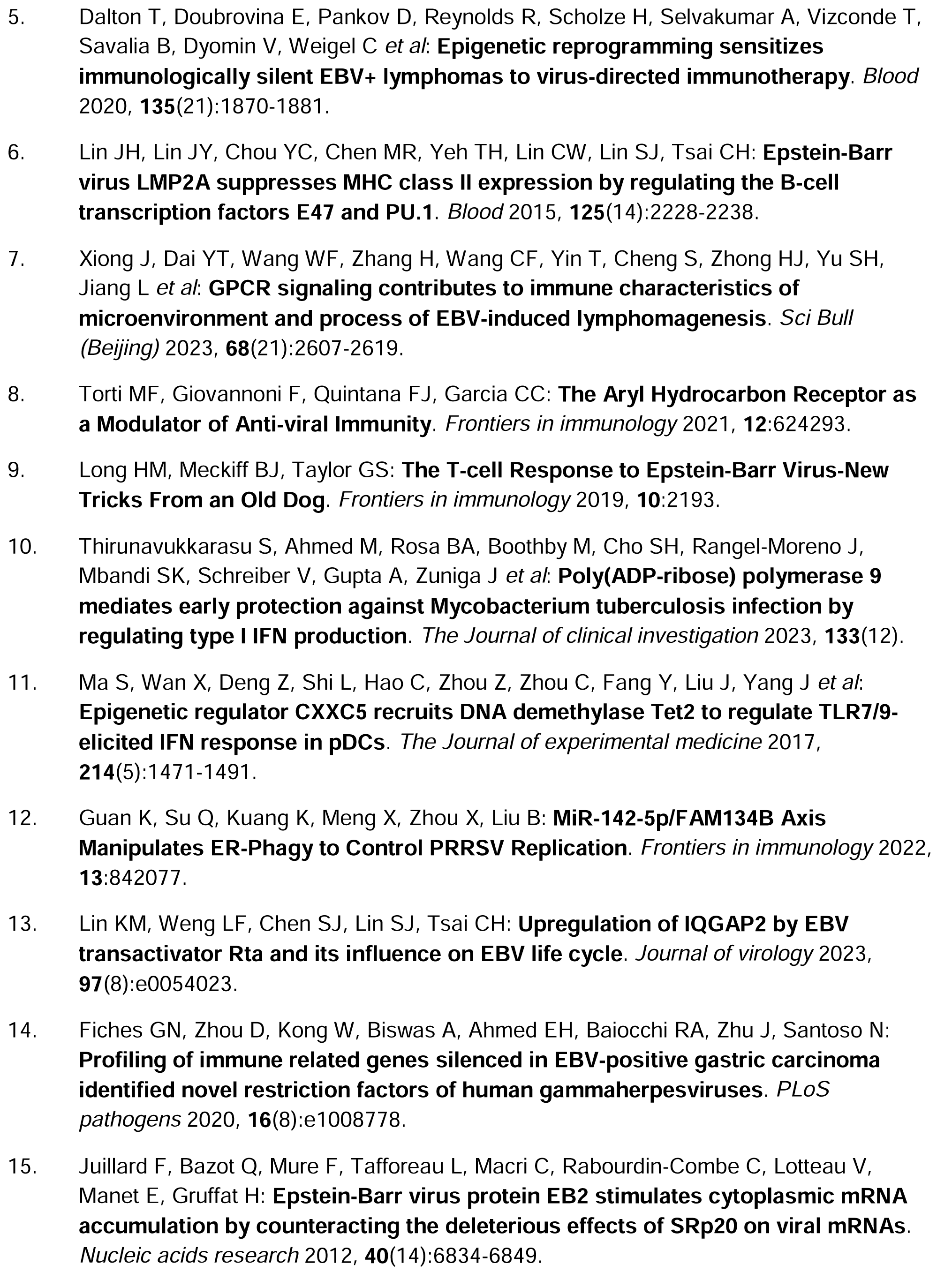
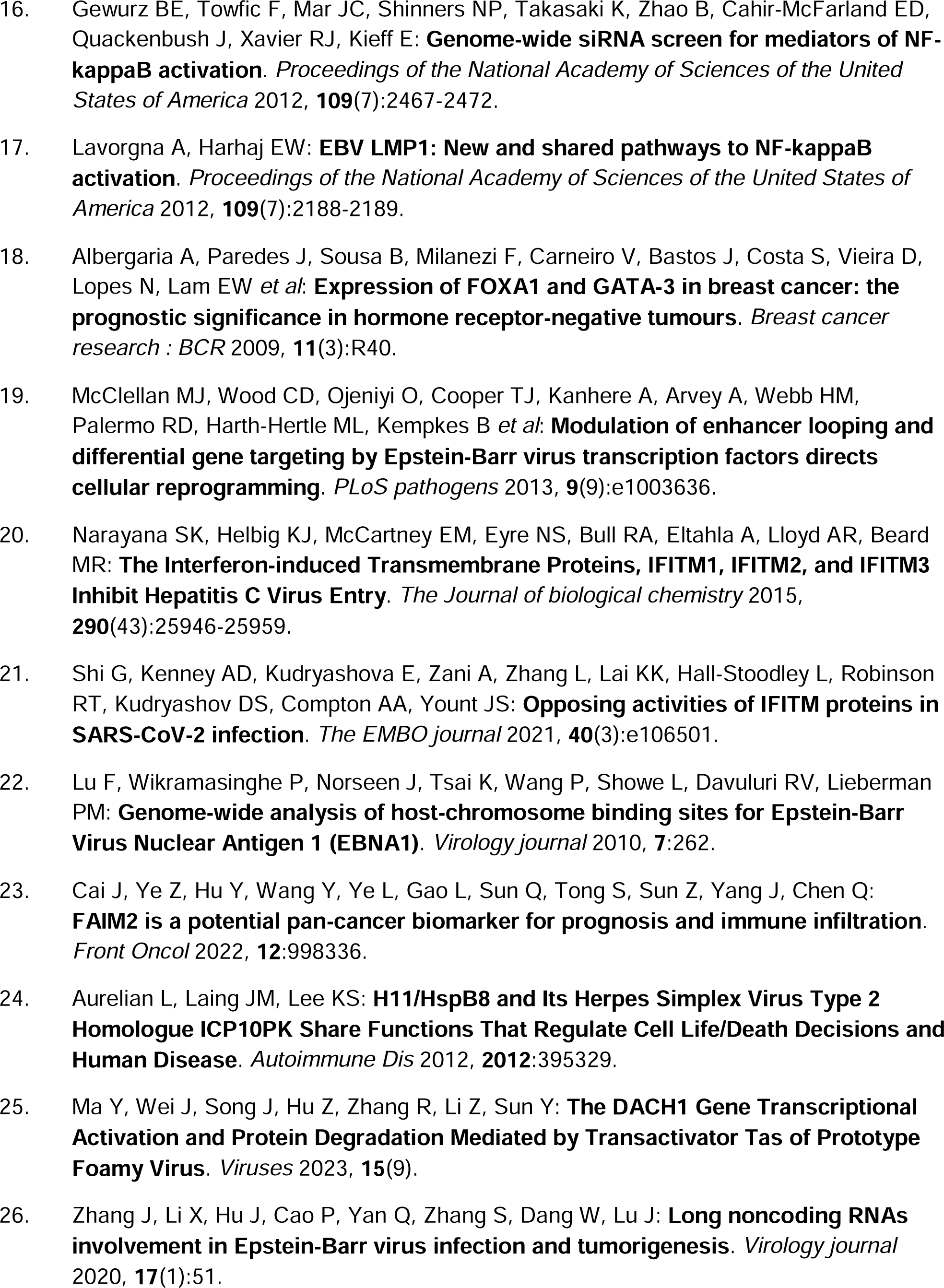
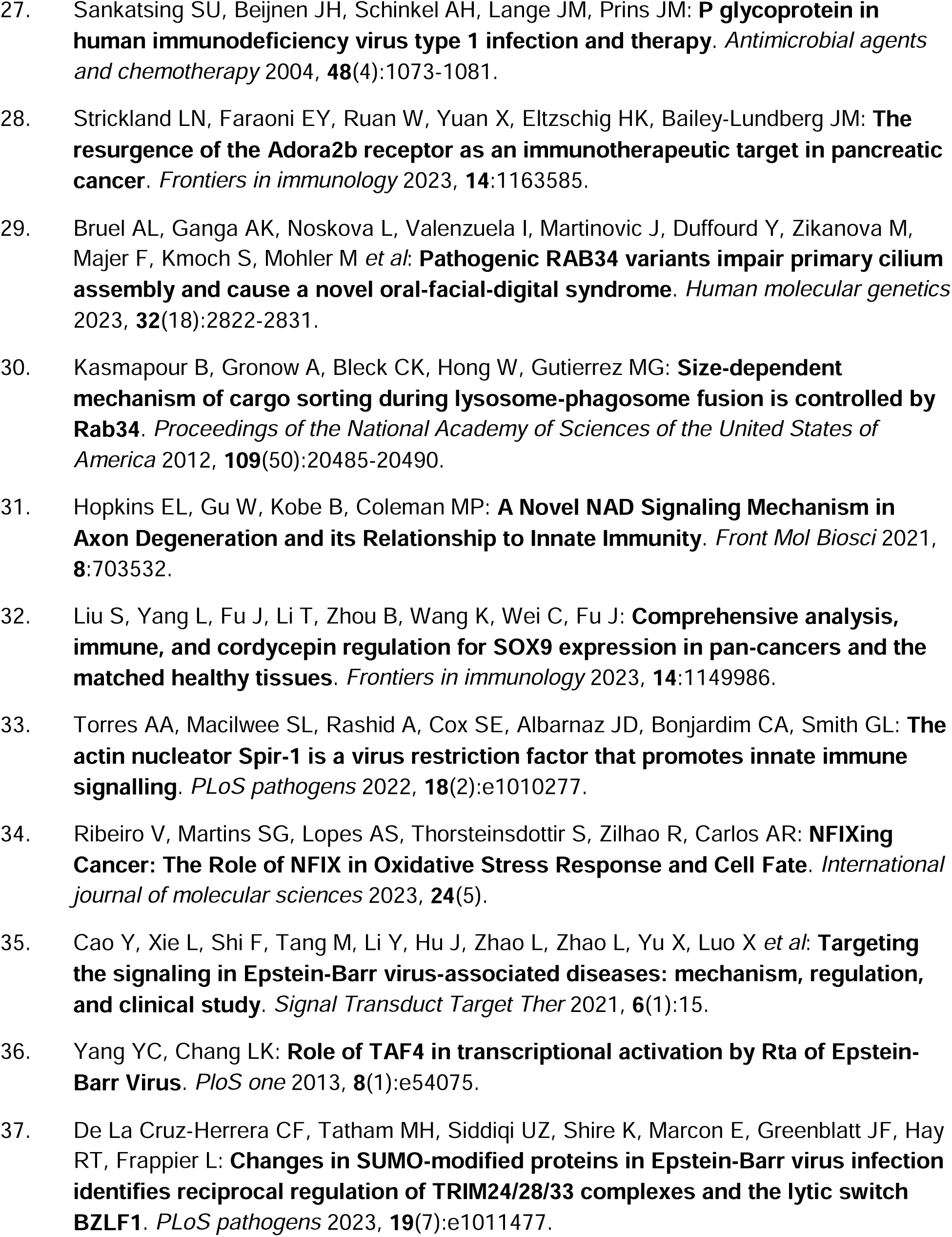
Hypermethylation in breast cancers inhibits genes that are essential for protection against EBV-related cancer (references listed in Table S2)

If EBV has a role in driving breast cancer, then one would expect that antiviral defense mechanisms would also be disabled. On chromosome 11, breast cancers and NPCs both had methylation clustered around genes that respond to interferons (65 to 161-fold enrichment, p=1.4e-4 to 9e-4). Methylation affects induced transmembrane proteins (IFITM1, IFITM-2, and IFITM-3), which cooperate in interferon-stimulated cells. IFITM3 fuses with incoming virus particles to send them to lysosomes [26]. Deficiencies in the breast cancer antiviral immune responses were related to defects caused by EBV infection in NPC (**Table 2**). This result shows that interference with antiviral defenses in breast cancer resembles the interference in an EBV mediated cancer.

### Chromosome 3p deletions in EBV cancers can create BRCA1/2-like deficits and cause the Warburg effect

I noticed that almost all NPC genomes (80% [27], 81-100% [28], 96% [29]) have deletions in at least one locus on chromosome 3p. These 3p deletions strongly associate with EBV: 100% of NPC with chromosome 3p deletions from a high-risk region of China were positive for EBV infection [28]. Precancerous lesions also lose heterozygosity (LOH) on chromosome 3p, implying chromosome 3p deletions are early events [28, 29]. To determine the effect of these deletions, known protein coding transcripts from reference genome sequence data were tabulated. Transcripts essential for HR and anti-EBV immune responses cluster together (**Fig. 5A**). During normal highfidelity DNA repair, DNA damage directs FANCD2 to nuclear foci where BRCA1 and BRCA2 proteins operate [30]. NPC deletions reported [31] remove FANCD2 from chromosome 3p, effectively disabling HR and allowing chromosome breaks to persist (supplementary Fig. 1). Loss of the nearby VHL tumor suppressor to the deletion further impairs HR repair [32]. These considerations show that EBV cancer loses protective mechanisms from chromosome 3p. These losses approximate inheriting a defective BRCA1 or BRCA2 gene because these genes depend on transcripts encoded on chromosome 3p (**supplementary Fig.1**).

**Fig. 5A.**
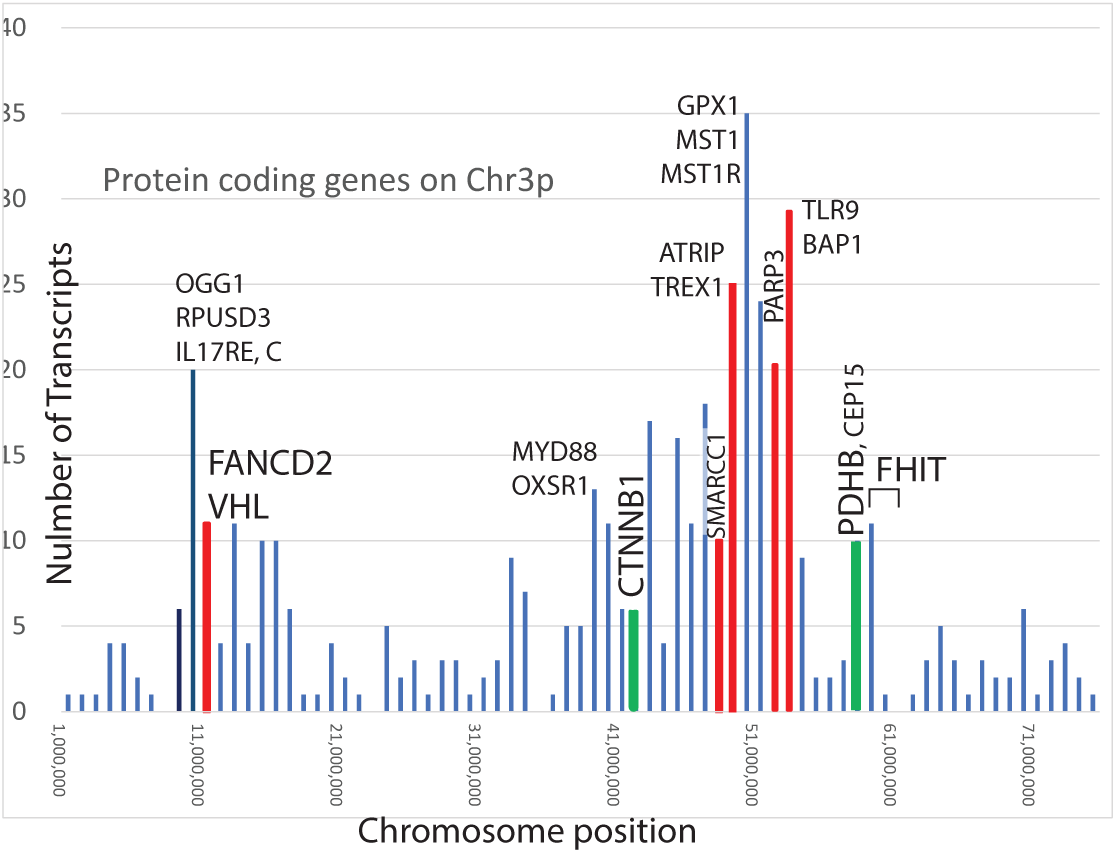
Selected protein-coding genes on chromosome 3p. Transcripts essential for HR are colored red, and transcripts necessary for oxidative phosphorylation are colored green.

#### Chromosome 3p deletions in breast cancers are similar to those that occur in NPC.

As in NPC, deletions affecting chromosome 3p are found in up to 87% of breast cancer cases, and nearly 20% of breast ductal tumors completely lose FANCD2 expression due to deletions on chromosome 3p [33–44]. The frequency and extent of 3p losses increase with the severity of pathological changes [45]. The fragile site FRA3B including FHIT is nearby [46] and is another source of frequent breakages on chromosome 3 (Fig 5A), with a potential connection to EBV infection [47].

Chromosome 3p includes the gene encoding the catalytic portion of pyruvate dehydrogenase (PDHB at chr3:58,427,630-58,433,832 ). EBV tumors without PDHB cannot metabolize pyruvate by oxidative phosphorylation [48, 49], so these tumors cannot use available oxygen to produce energy most efficiently from glucose (the Warburg effect). TCGA data finds deletions, including PDHB occur in about 30% of breast cancers when comparing results from multiple datasets (**Fig 5B**).

**Fig. 5B.**
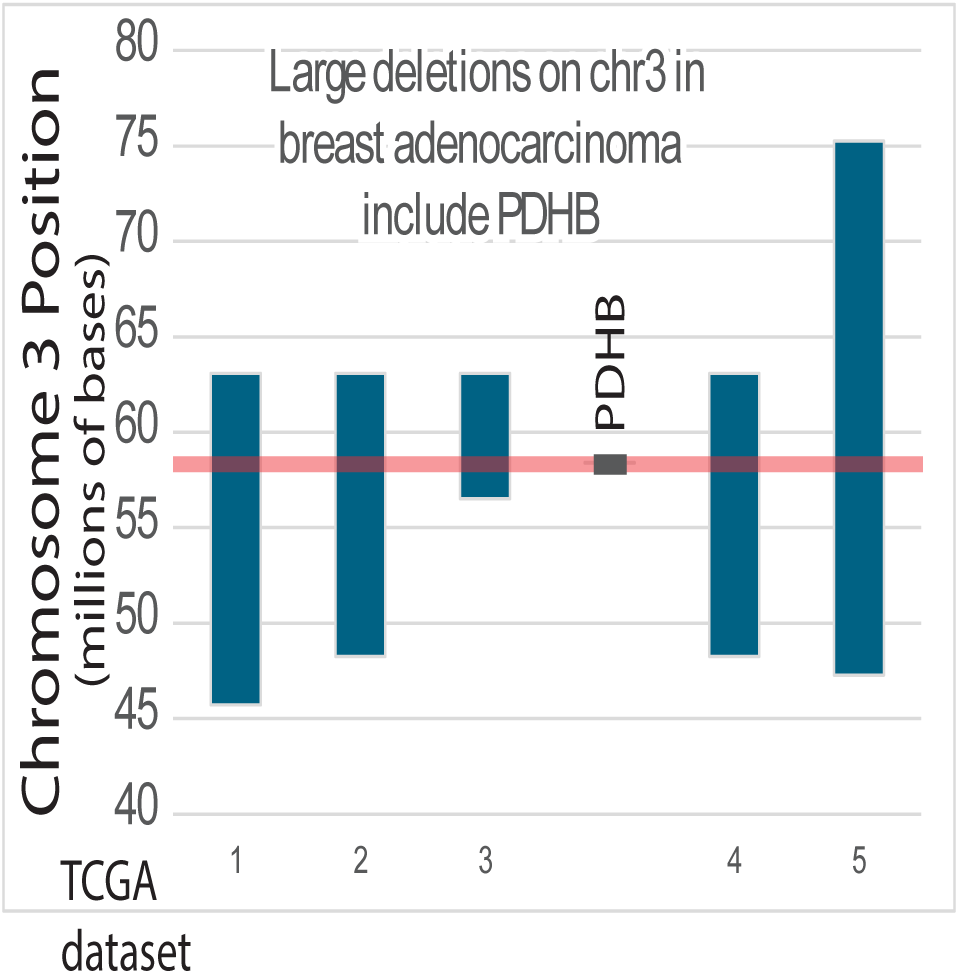
Deletions on chromosome 3p from five TCGA datasets include the pyruvate dehydrogenase catalytic subunit (PDHB).

To test for additional changes in chromosome 3p in breast cancer that might be responsible for the Warburg effect, I plotted frequencies of known short deletions on chromosome 3p in COSMIC data from 476 breast cancers (**Fig. 5C**). These deletions are typically caused by AID [50], which EBV induces prior to lymphomas [51, 52]. The deletions (**Fig 5C**) mainly target transcript variants of CTNNB1, which then deregulate WNT/CTNNB1/TCFL2/PDHB signaling, causing the Warburg effect. EBV-positive gastric cancer targets the same pathway [8]. Furthermore, breast cancers also have hypermethylation of PDHB. Because glucose metabolism is complex and tightly controlled, other genetic abnormalities probably contribute to the Warburg effect. A breast cancer breakpoint approximately the same as an NPC breakpoint is only 6743 base pairs from the PDHX gene, a different subunit of the pyruvate dehydrogenase complex. PDHX is within a fragile site on chromosome 11 [46].

**Fig 5C.**
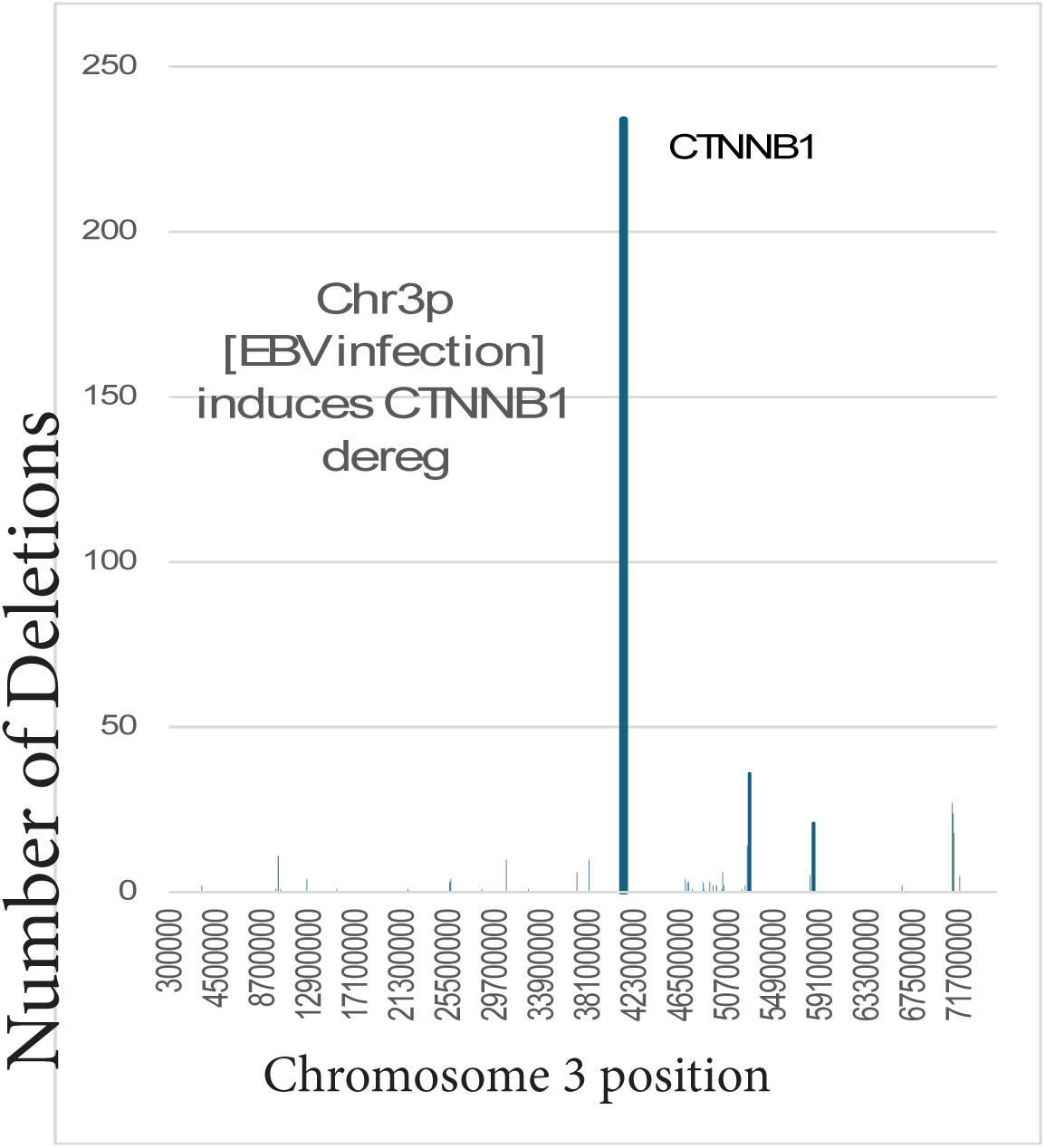
The short deletions on chromosome 3p in breast cancers listed in the COSMIC database focus on CTNNB1. This damage should deregulate PDHB.

The microenvironment of early breast cancer strongly selects for the Warburg phenotype [53], and almost all breast cancers have it. The Warburg phenotype associates with progression in triple-negative breast cancer [49]. These results show that EBV infection can contribute to both defective DNA repair and metabolic reprogramming in breast cancer.

### Breakpoints in EBV-associated NPC and breast cancer occur at similar positions

The HR deficits observed in NPC are like those in hereditary breast cancer, so it is possible that breakpoints are also similar, especially those associated with translocations and oncogene amplification. To test this possibility, every breast cancer translocation breakpoint in a dataset from 780 breast cancers [54] was compared to all breakpoints on corresponding chromosomes from 70 NPC’s [16]. The results revealed that both breast cancer and NPC had many similar breakpoints on all chromosomes (**Fig. 6A**), but chromosome 11 had the largest number of similar breakpoint positions. On some chromosomes, Mann-Whitney testing gave P values >0.05 that could not rule out identical overall breakpoint distributions in NPC and breast cancer (**Fig. 6A**).

**Fig. 6A.**
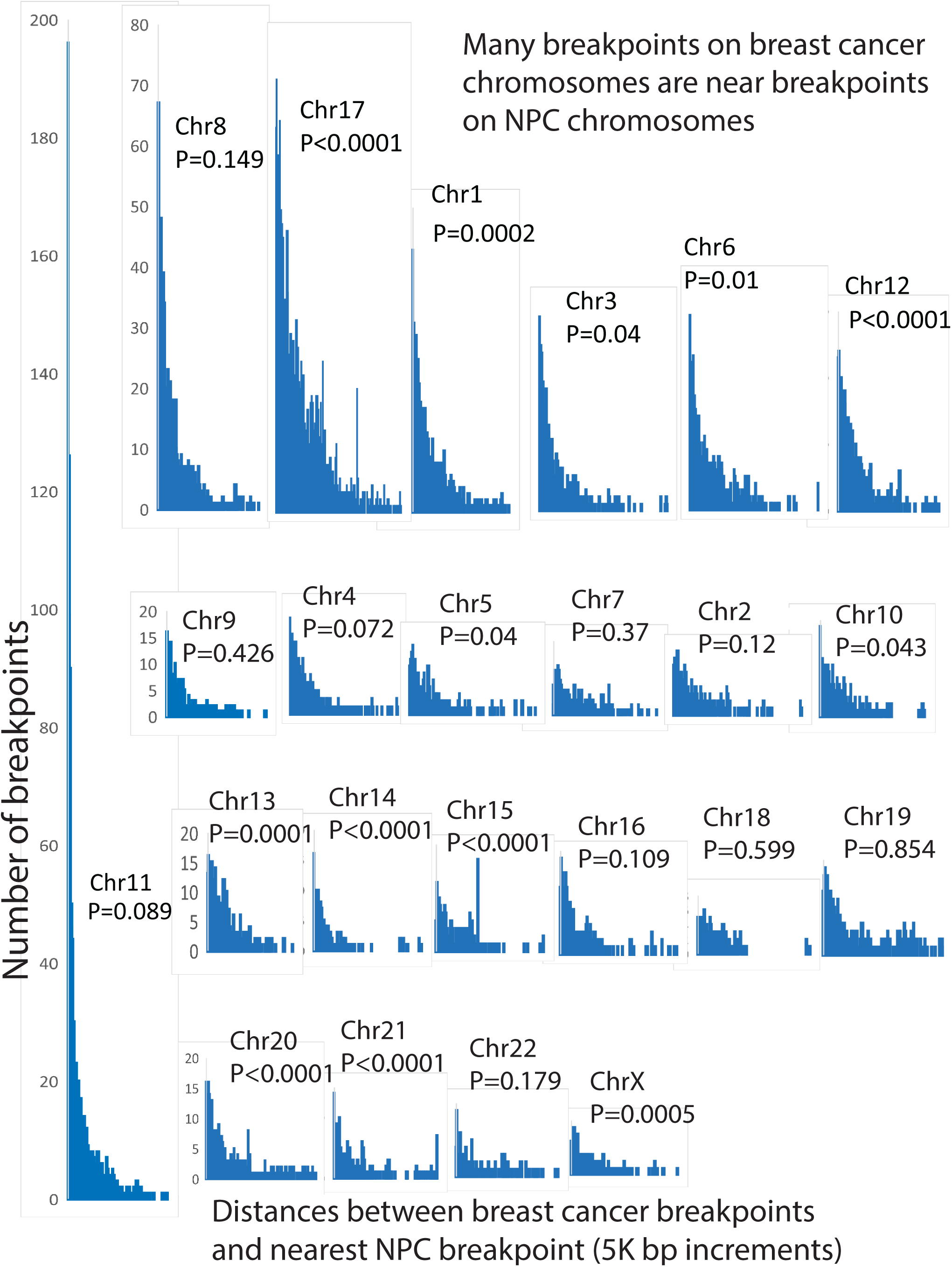
Distances separating the nearest breakpoints in breast cancer from breakpoints in the EBV cancer NPC. P values are from Mann-Whitney calculations and represent probabilities that the distributions are the same. P>0.05 indicates that the possibility that the breakpoint distributions are identical cannot be excluded.

To verify this result, breakpoint positions on chromosome 11 in NPC and breast cancer were compared further (**Fig 6B**). Breast cancers and NPC’s shared a large cluster of breakpoints telomeric to the centromere between 55 - 80 million bps. This region includes the CCND1 gene, which is known to be amplified in NPC [16] and is a site for breast cancer translocations and oncogene amplifications [54]. Further comparisons revealed the frequencies of breakpoint positions in breast cancers and NPC were similar on chromosomes 8, 11 and 17 (**Fig. 6B**).

**Fig. 6B.**
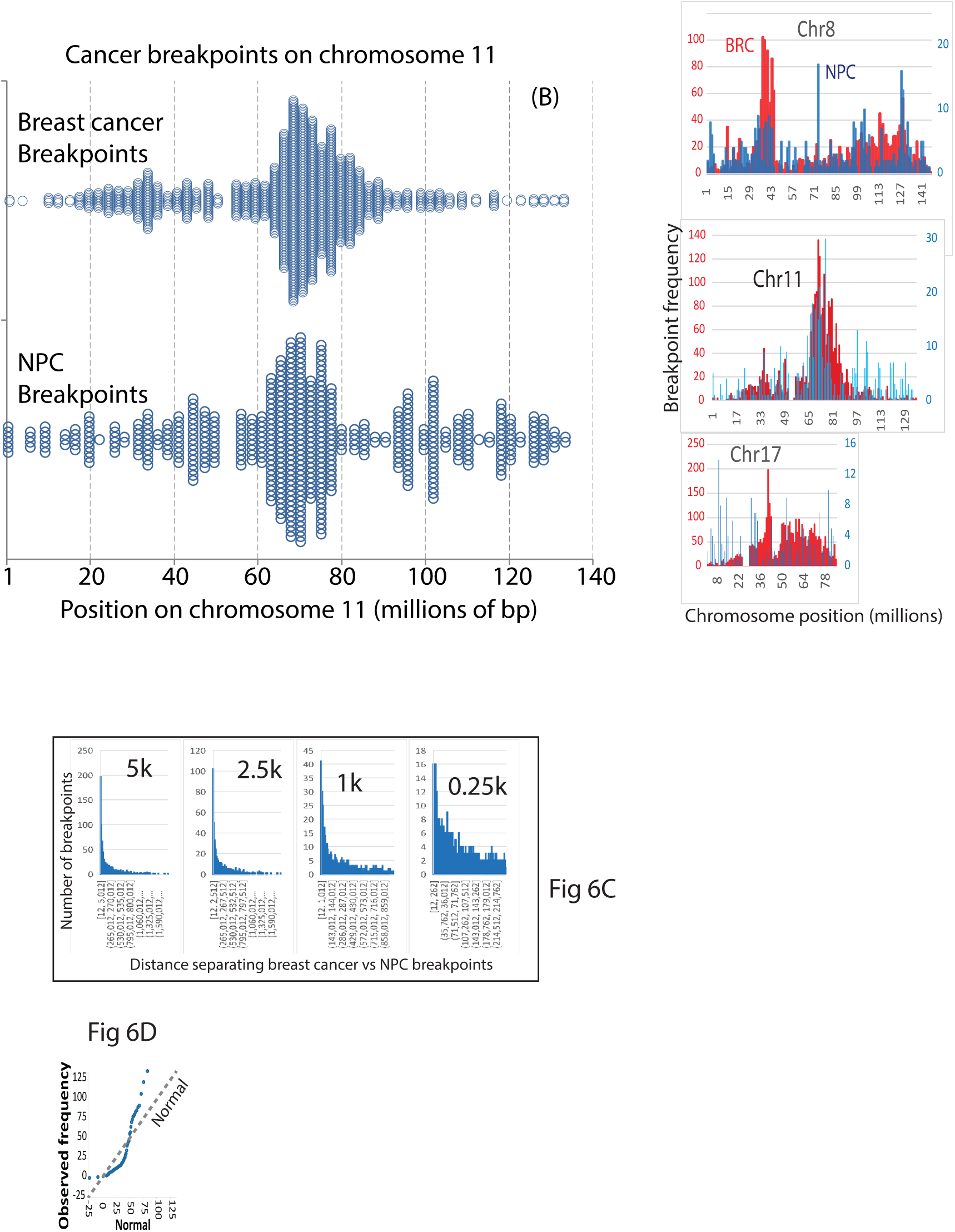
Plotting the number of breakpoints or the frequencies of breakpoints vs. chromosome positions in breast cancer and NPC illustrates chromosome positions where breakpoints are near each other. Fig. 6C. The relationship between some NPC and breast cancer breakpoints on chromosome 11 is evident down to a window of 250 base pairs. Fig 6D. Tests for normality of the distribution of nearest breakpoints show that the values are unlikely to be normally distributed.

The numbers of breakpoints in breast cancer that agreed with NPC depended on the window size, but smaller windows did not change the conclusions. For chromosome 11, the use of a window of 2.5k, 1k, 0.5k, or 0.25k instead of either 5k or 10k produced fewer breakpoints that approximately coincided, but did not change the result that some breast cancer breakpoint were near NPC breakpoints (**Fig. 6C**). Normality tests indicated that breakpoint positions in breast cancer, NPC, or their frequency distributions on chromosome 11 were unlikely to represent normal distributions (**Fig 6D**). Based on the above results, breakpoints in an EBV mediated cancer occur near some of the same chromosome translocations that drive breast cancer.

### Breakpoints in the EBV cancer BL are also similar to breast cancer breakpoints

To determine whether the resemblance between breakpoints in EBV cancer and breast cancer depends on whether the tissue is epithelial or lymphoid, I compared published copy number and structural variants (CNV and SV) in chromosomes from 94 EBV-positive children with BL [15] to translocation breakpoints in 780 breast cancers [54]. Many breast cancers have chromosome breaks closely related to breakpoints in EBV-positive BL (**Fig. 7A**). Polynomial regression analysis found that breakpoint numbers on BL chromosomes predicted breakpoint numbers in breast cancer, albeit according to a polynomial curve (**Fig 7B**). Specifically, the coefficient of determination (r-squared) value suggested that the breakpoint numbers in EBV-positive BL could account for approximately 48.6% of the variability in breast cancer breakpoint numbers. The polynomial curve suggests that the distance between BL and breast cancer breakpoints somehow affects the total numbers of breakpoints. In summary, the relationship between breast cancer breakpoints in BL, an EBV cancer in lymphoid tissue, is consistent with EBV influencing or causing some of the chromosome breakpoints.

**Fig. 7A.**
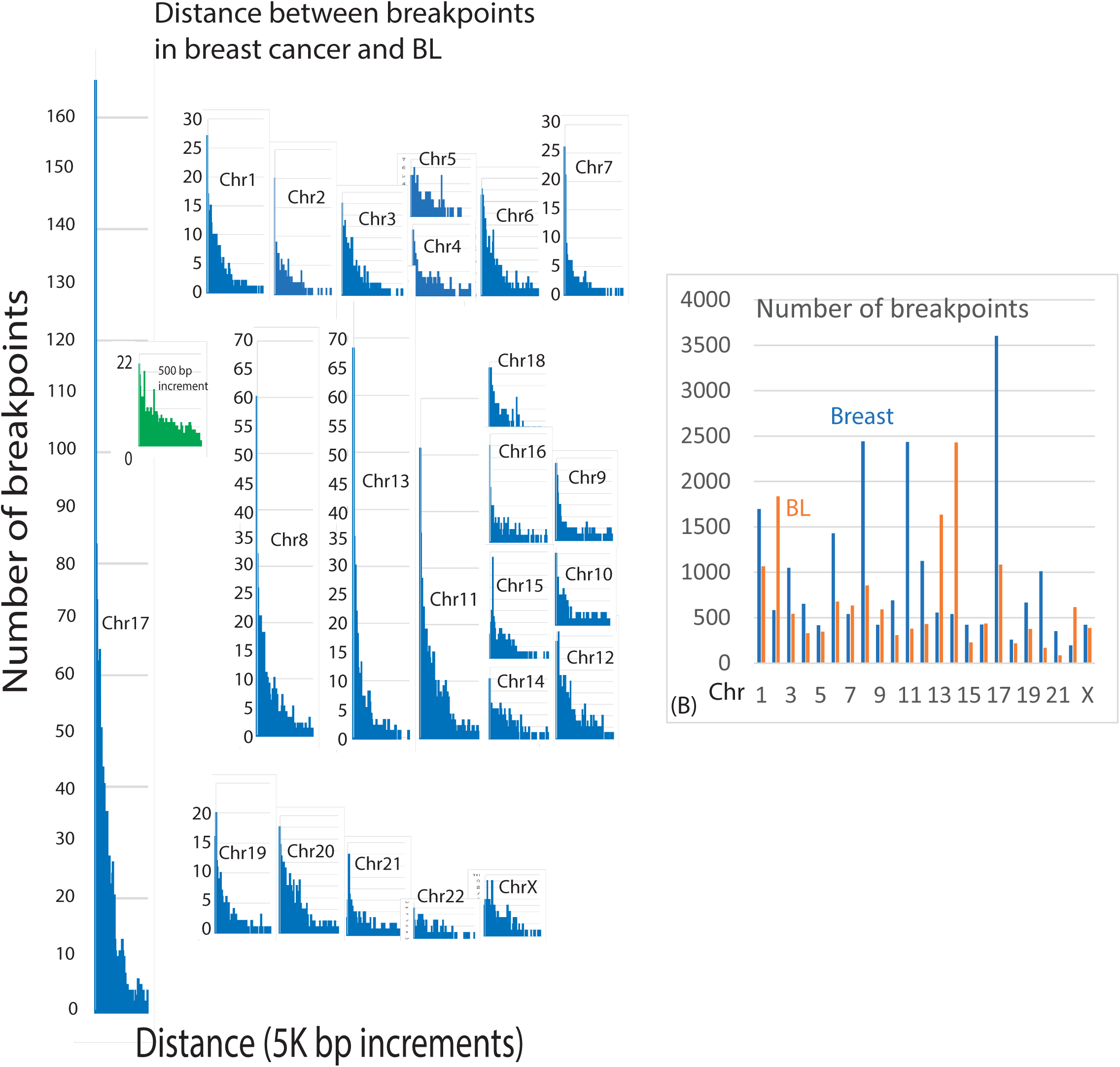
Distances between breakpoints in breast cancers nearest those in EBV-positive BL. The proximity of breast cancer and BL breakpoints on chromosome 17 is still apparent down to a value of about 500 base pairs. Fig. 7B. Numbers of breakpoints for each chromosome in breast cancer vs. BL. The numbers are statistically related by polynomial regression analysis.

### Breast cancer translocations and focal amplifications are related to somatic hypermutations in EBV cancers

Identifying a plausible mechanism that EBV could use to drive breast cancer began by reviewing the relationship between EBV and AID. EBV-positive BL has increased AID activity in vivo [15] and NPC also has AID footprints [12]. AID converts cytosine to uracil that mimics dT during DNA replication. Uracil causes mutations if it pairs with G because the U:G mismatch triggers error-prone DNA repair. Then uracil DNA glycosylase and/or DNA mismatch repair enzymes produce local mutations and double-strand breaks as they remove DNA mismatches [55]. These events cause the DNA double-strand breaks that produce c-myc/IgH translocations characteristic of BL. If EBV is involved in breast cancer, then breakpoints at breast cancer translocations and focal amplifications could also correspond to positions where AID deregulation causes SHM. The MYC locus is a prototypical example of translocations and amplifications accompanying AID deregulation.

The first test of this hypothesis was to determine whether EBV cancers and breast cancers have similar breakpoints on chromosome 8 around the MYC gene locus. The results revealed that the distance between NPC and BL breakpoints plotted against the number of breakpoints was approximately linear up to about 11,000 bp (**Fig 8A**). The first 11,000 base pair interval included about 75 breakpoints (**Fig. 8B**). This result implies that EBV induces a set of related breakpoints in NPC and BL. Consistent with this conclusion, various breast cancers had over 400 breakpoints near the MYC locus (**Fig. 8C**). These results show that MYC locus breakpoints characteristic of EBV cancers are near breakpoints in breast cancer.

**Fig 8A.**
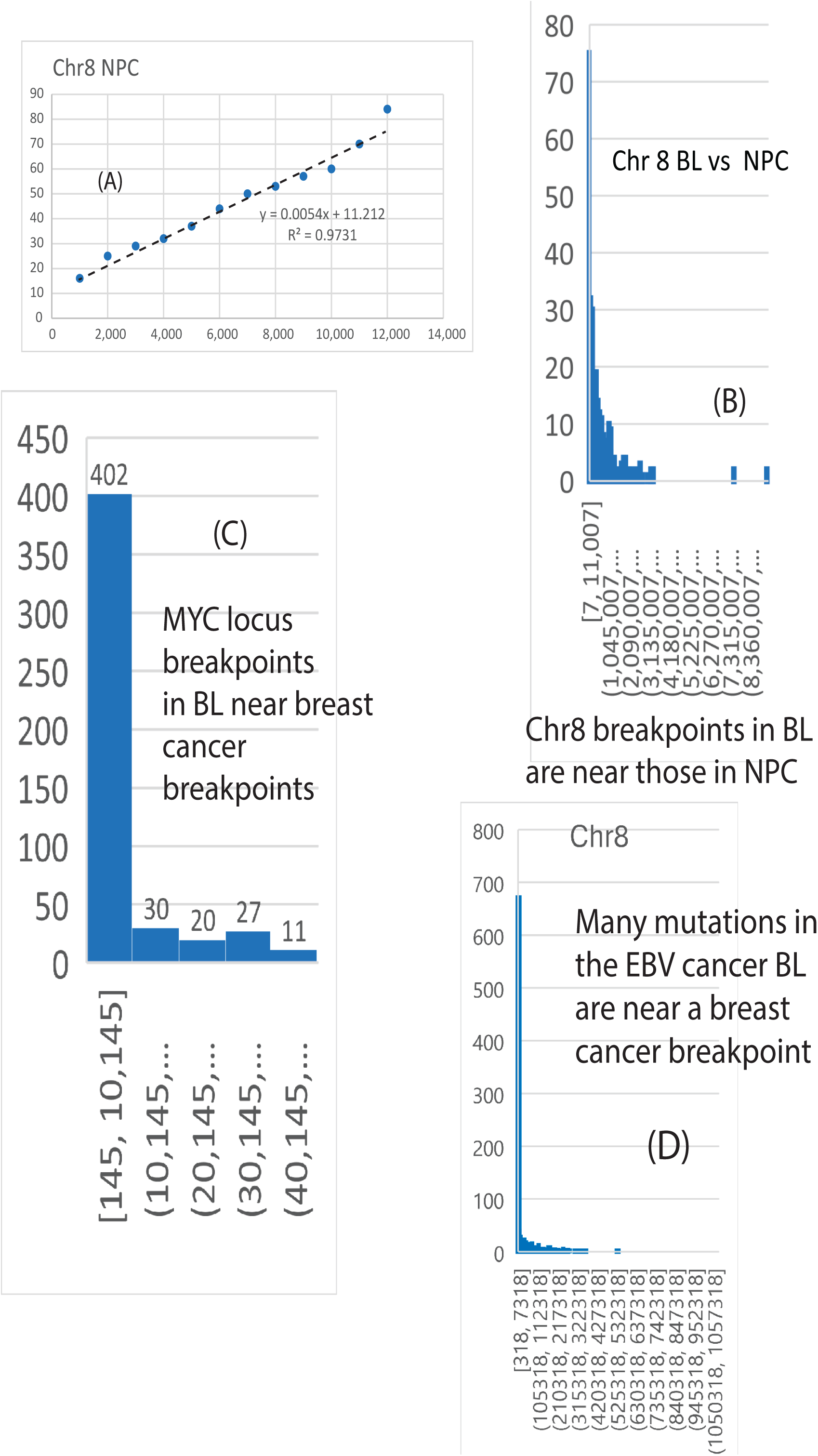
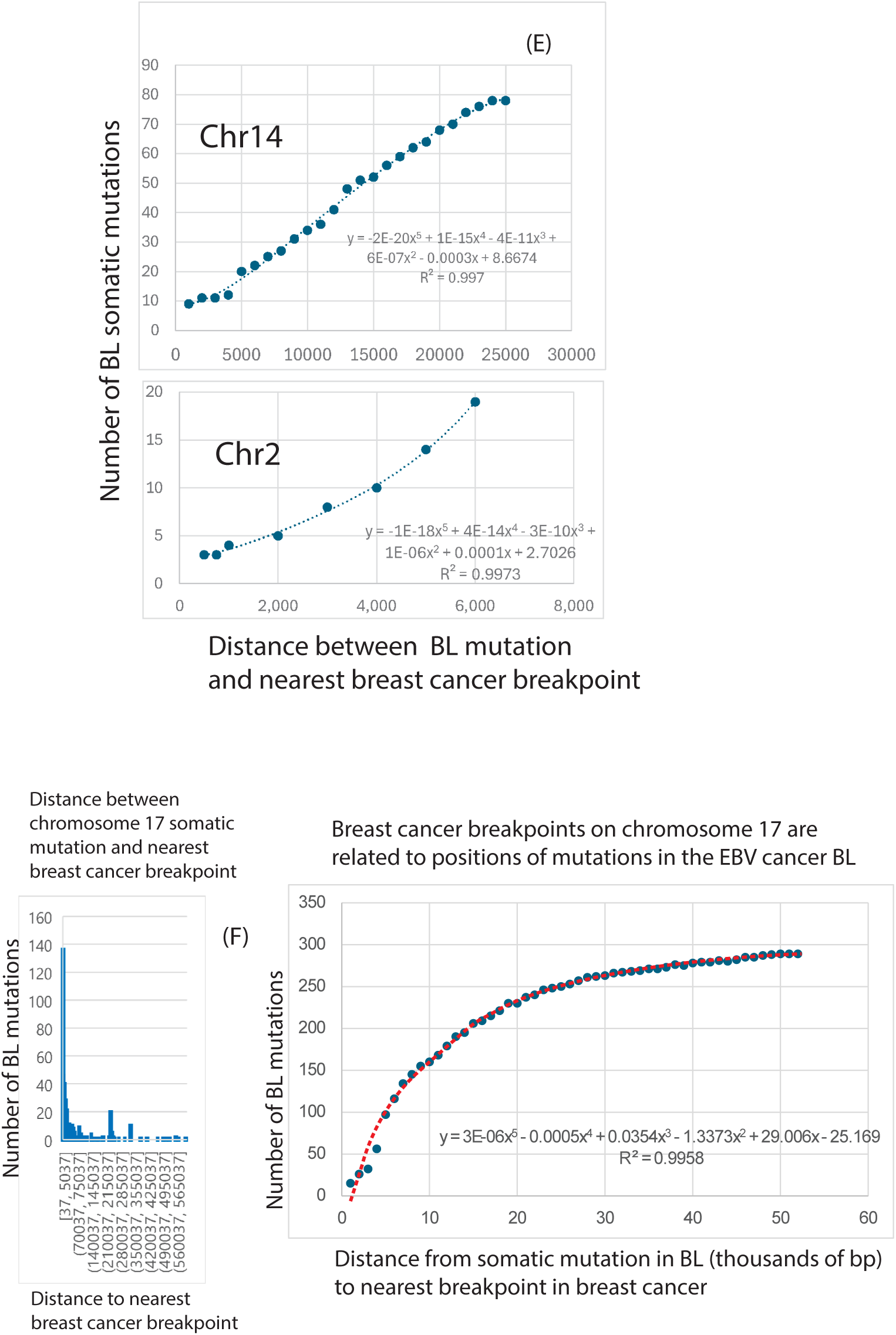
Breakpoints on chromosome 8 in BL are near those in NPC. The numbers of breakpoints increase linearly as the window size increases up to about 11,000 bp. (B). About 75 breakpoints in NPC are near a BL breakpoint within the 11,000 bp window. (C). Various breast cancers had over 400 breakpoints near MYC locus breakpoints on chromosome 8 in BL. (D). Nearly 700 BL somatic mutations are within 7318 bp of a breast cancer breakpoint on chromosome 8. (E). Chromosome 14 and chromosome 2 translocation partners to chromosome 8 also show associations between AID induced SHMs in BL and breast cancer breakpoints. (F). Chromosome 17 has the largest number of breast cancer breakpoints related to BL. Nearly 140 BL mutations on chromosome 17 are within 5k bp of a breast cancer breakpoint. The overall distribution of BL mutations has a complex polynomial relationship to breast cancer breakpoints. This shows that the effect of somatic mutation on breast cancer breakpoints changes at different separation distances.

EBV is capable of inducing AID [56–58]. If EBV mediates AID deregulation in breast cancer, then SHM in EBV related cancers might also be near translocation and focal amplification sites. To test this hypothesis, the positions of SHM in BL on chromosome 8 were compared to breast cancer breakpoints. The results confirmed that the majority of EBV associated SHMs in BL on chromosome 8 (685/1221) were near a breast cancer breakpoint (**Fig 8D**). In translocations characteristic of BL, chromosome 14 and chromosome 2 are significant partners with MYC, so BL somatic mutations were then compared to breast cancer breakpoints on chromosomes 14 and 2. Breast cancer breakpoints were again related to positions of SHM (**Fig 8E**). Chromosome 17 had the largest number of breakpoints related to BL (**Fig 7A**) and was used to further test for a relationship between EBV-associated SHM in BL and breast cancer breakpoints. Nearly 140 positions of somatic hypermutation in BL were within 5k bps of a breast cancer breakpoint (**Fig. 8E**) and the relationship fit a complex polynomial (**Fig 8F**). These results establish that SHM in EBV cancers, induced by AID deregulation, occurs near breakpoints in breast cancer and includes the characteristic MYC locus.

Estrogen has been proposed as directly generating initial translocations in estrogen receptor targets in human breast cancer [54]. However, BL in mostly male pediatric patients has breakpoints [59] like those in breast cancer so BL data conflicts with the role of estrogen in initiating translocations. Instead, transcription at elements that respond to estrogen-receptor complexes requires topoisomerase. Topoisomerase causes transient DNA breaks that are normally repaired by HR-proteins [60]. In breast cancer, topoisomerase is a strong predictor for amplification at translocation boundaries [54]. EBV-infected cells also lose normal control of aromatase, an enzyme which converts estrogen precursor into estrogen. Transcription in response to high estrogen levels induces topoisomerase mediated DNA breaks but herpesviruses can also hijack topoisomerase [61]. The expression of ZEBRA (the EBV lytic switch in NPC) increased levels of aromatase and ER-α [62]. According to this hypothesis, it is EBV mediated deregulation of AID, estrogen production from EBV control of aromatase, and topoisomerase that drives breast cancer translocations. If so, then the distribution of breakpoints and focal abnormalities in breast cancers should be relatively independent of ER status.

To test these explanations, I compared 10736 breakpoints in 271 ER-negative breast cancer cases to 12242 breakpoints in 473 ER-positive breast cancer cases. The results revealed hundreds of breakpoints near each other on some chromosomes (**Fig. 9A**). On chromosome 17, 29 breakpoints in ER-negative breast cancers were within ten bp of a breakpoint in ER-positive breast cancer. In contrast, a random breakpoint control had only 0-3 breakpoints within 10 bp of an ER positive breakpoint. Chromosome 8 had over 70 breakpoints within 1500 bp of one another (**Fig. 9A**, right). Chromosome 11 had about 130 breakpoints within 5k bp of one another (**Fig. 9B**) and their frequency distributions had a region of local overlap (**Fig 9C**). The proximal breakpoints focused on chromosomes 17, 8, and 11, but all chromosomes had them (**Fig 9A**). The distributions of breakpoints in ER-positive vs. ER-negative breast cancer could not be confidently distinguished (P>0.05).

**Fig. 9.**
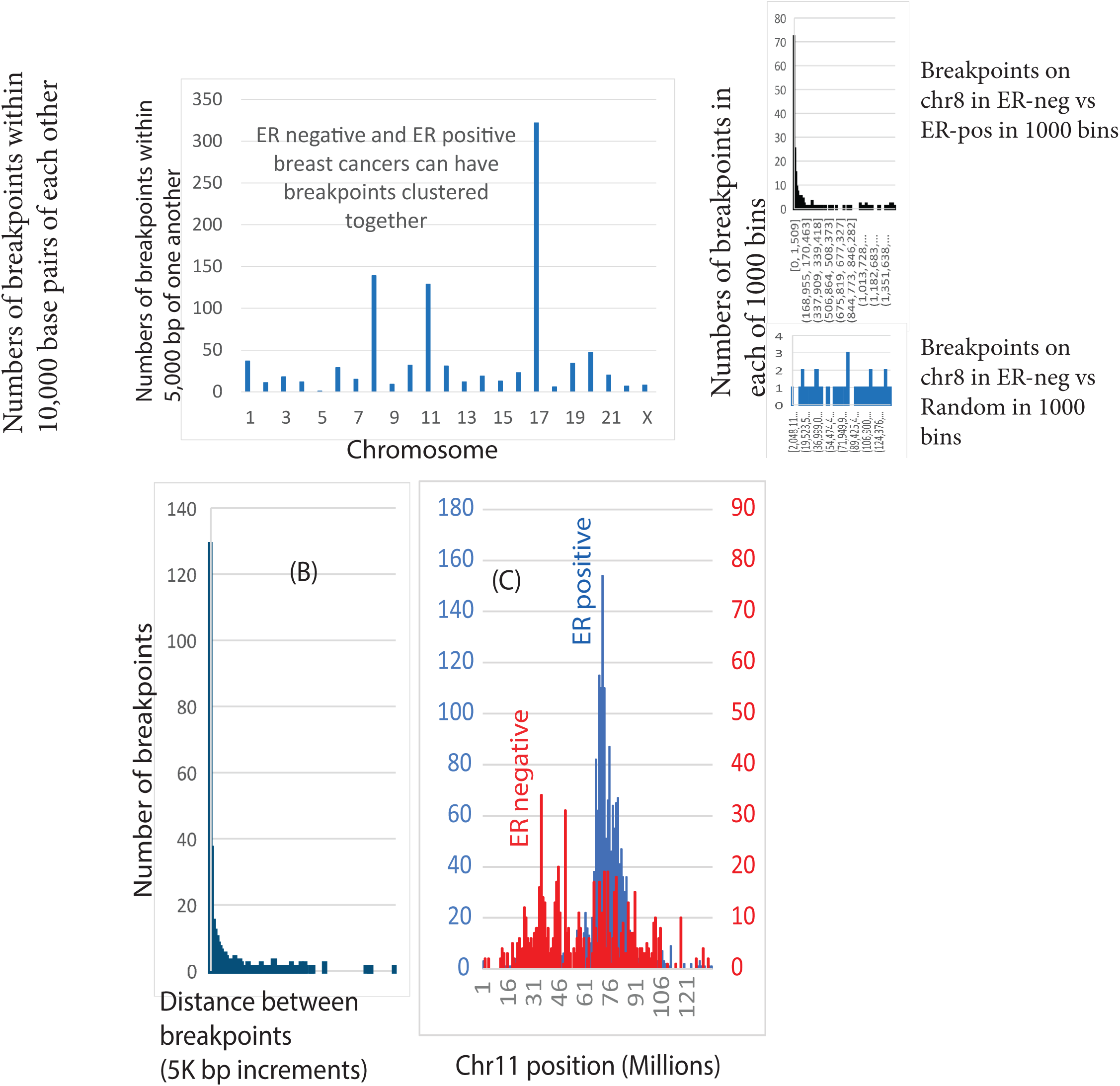
ER status in breast cancers is not sufficient to predict breakpoint positions because ER-negative breakpoints could be near ER-positive breakpoints. Fig 9A. Summary of the numbers of breakpoints within 10,000 base pairs of each other in ER-positive vs ER-negative breast cancers. At the right of Fig. 9A is a comparison of the breakpoint distributions between ER negatives vs. ER-positive samples (top) vs. a comparison between ER negative samples and a random distribution (bottom). Both comparisons are from chromosome 8 divided into 1000 bins. Fig 9B. Abount 130 breakpoints on chromosome 11 in ER-positive and ER-negative breast cancers are within 5K bp of each other. Fig. 9C. Comparisons of the frequency distributions of breakpoints in ER-positive vs ER-negative breast cancers on chromosome 11.

The effect of estrogen on translocations and focal oncogene amplifications in breast cancer [54] is undeniable, but the above evidence shows that ER status does not determine the location of chromosome breaks. The results instead show EBV fits breast cancer and BL data better than differences in ER status because of AID deregulation, inappropriate estrogen production from aromatase with herpes virus mediated deregulation of topoisomerase.

### Many chromosome breakpoints occur at approximately the same positions in HR-deficient and HR-proficient breast cancers

If EBV creates deficits in HR, then breakpoints in HR-deficient (HRD) BRCA1/2-type breast cancers should resemble those in HR-proficient (HRP) breast cancers. To test this hypothesis, I compared the breakpoint positions in HRD vs. HRP breast cancers. The exact coordinates of all breakpoints in HRP vs HRD breast cancers were then compared to measure this apparent similarity more precisely (**Fig. 10**). All chromosomes with the possible exception of chromosome 22 showed some breakpoints on HRP breast cancers that were near (within 5000 bp) those in HRD breast cancers. On chromosome 17, >180 breakpoints were within 5000 bp of one another and 50 breakpoints were within 50 bp. For chromosomes 3, 6, 19, 22, and X, the Mann-Whitney test could not exclude the possibility that the entire distribution of breakpoints was the same (**Table 3**).

**Fig. 10.**
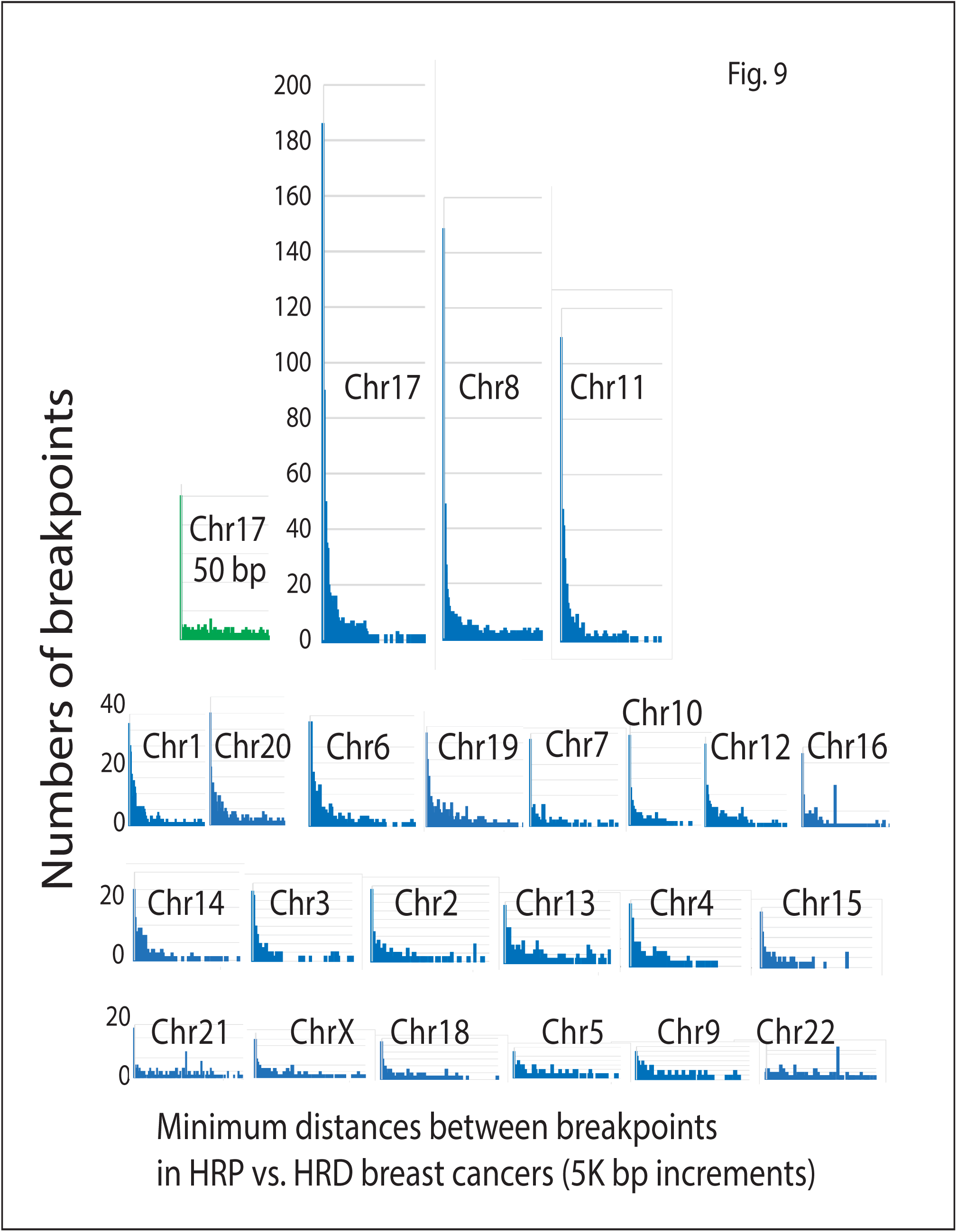
Comparisons of breakpoint distances between breast cancers that are HR proficient vs HR deficient (Increments of 5K bp). The first peak is the numbers of breakpoints within 5k bp and is the largest for all chromosomes except chromosome 22.

**Table 3.**
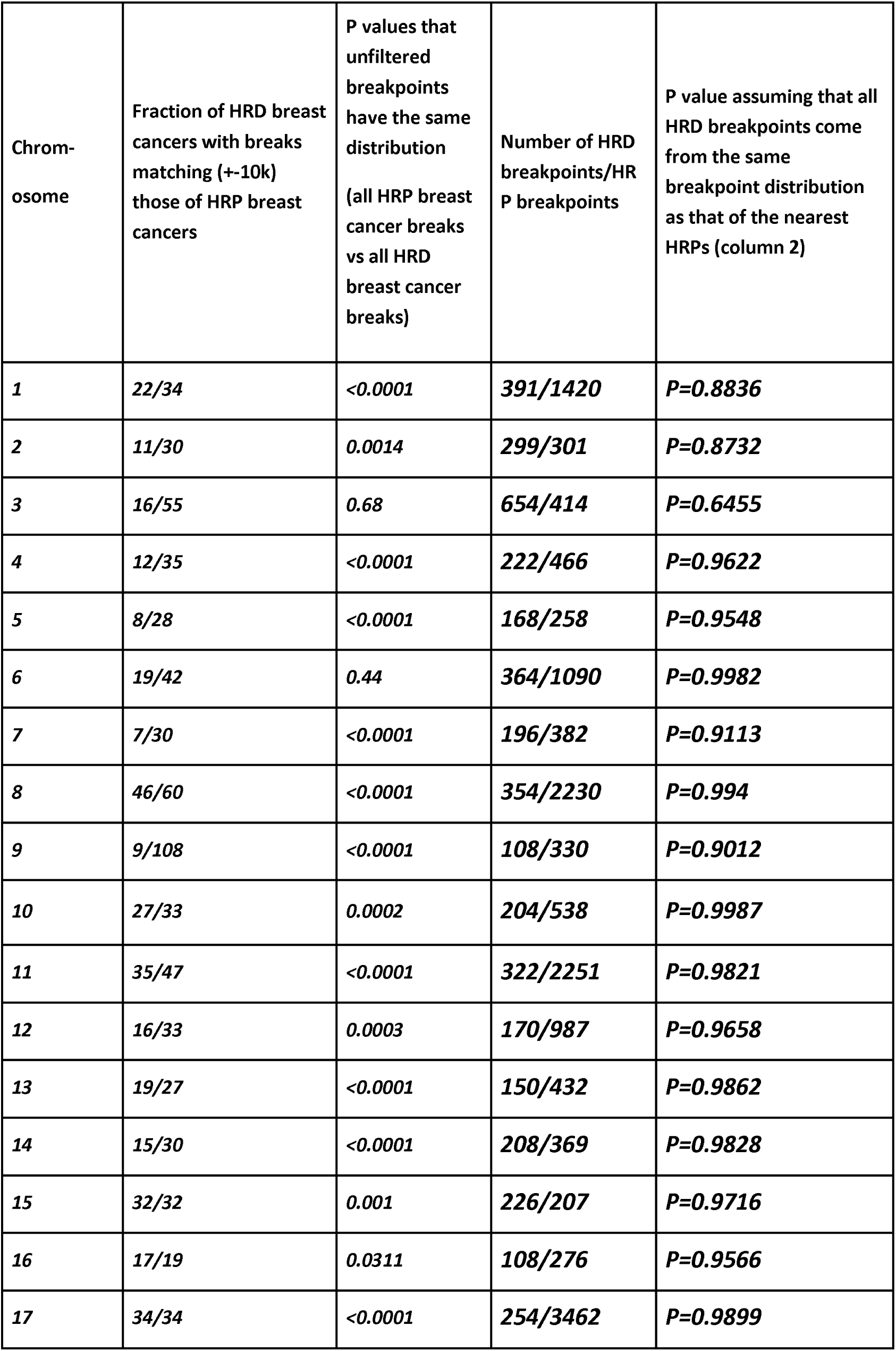
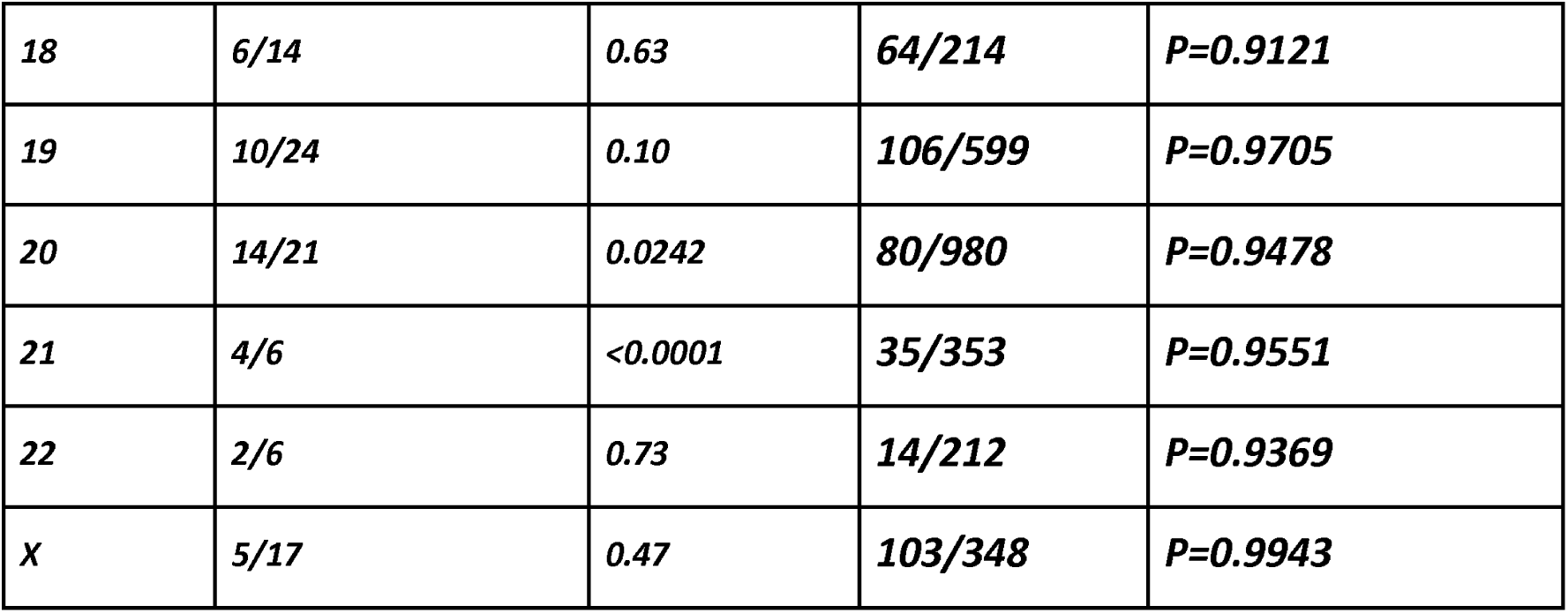
Relationships of breakpoints in HRD breast cancers to those in HRP breast cancers.

Next, the idea was tested that there are local areas of similarity even when the overall distributions of breakpoints differs. Local breakpoint distributions in HRD breast cancers matched local breakpoint distributions in HRP breast cancers in at least some chromosome sections, even if the overall distributions differed (**Table 3** and **Fig. 10**). The breakpoints in HRD breast cancer nearest those in HPR breast cancers probably follow the same distribution. Based on the above statistical comparisons, many breakpoint positions in HRD and HRP breast cancers are similar. These results are consistent with the idea that EBV inhibits HR to create a deficit in HRP breast cancers equivalent to hereditary pathogenic BRCA1 or BRCA2 mutations (HRD breast cancers).

### Breakpoints in EBV-positive BL genomes approximate the ability of BRCA1 or BRCA2 mutation to block a cancer suppressive mechanism

To test the idea that EBV can mimic hereditary BRCA gene mutations, I assessed breakpoint positions in EBV-positive BL in the context of well-known genes required for HR [63] or identified as HR genes from KEGG pathway analysis. BL breakpoints were found to affect HR-h2related genes mainly on chromosomes 1, 11, and 17 (**Fig. 11**). The most frequently targeted gene was H2AX, which is located on chromosome 11. On chromosome 13, BRCA2 was affected in 3 BL samples, and other breakpoints clustered near BRCA1 on chromosome 17. Although the data are limited, this finding related to HR damage echoes the results in NPC, showing that EBV-related cancers cause damage to genes associated with hereditary breast cancer risk.

**Fig 11.**
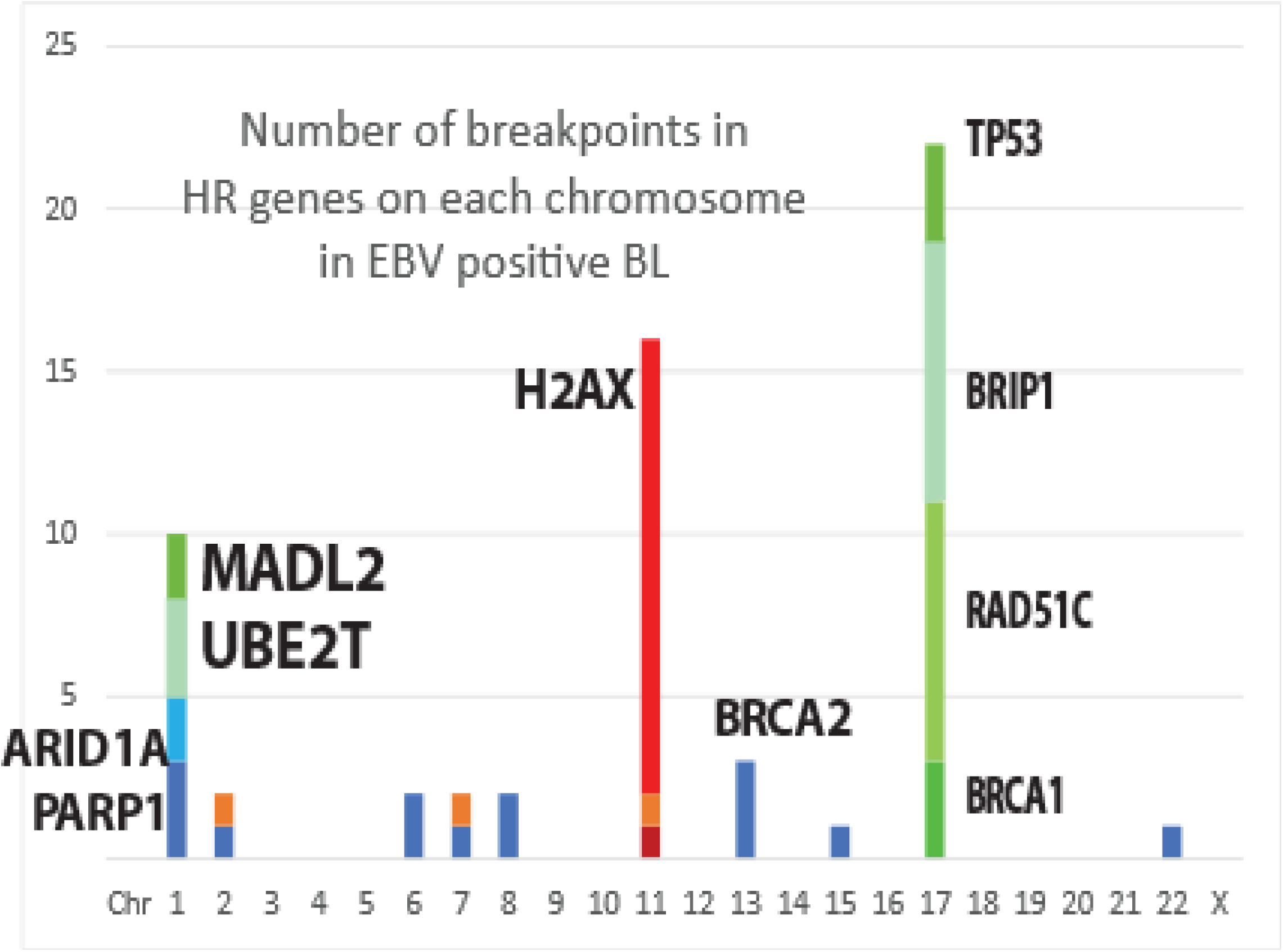
Breakpoints in EBV-positive BL affect genes essential for HR. A minimal list of these HR genes is compiled for each chromosome and indicated by different colored bars.

### Alternate Explanations

### The binding of the viral tethering protein EBNA1 binding to a DNA consensus sequence does not initiate chromosome breakage in breast cancers

An alternative explanation for EBV-related carcinogenesis involves the docking of EBNA1 virus tethering protein at specific EBV binding sequences clustered on chromosome 11 [64]. The host binding sequences consist of repeating imperfect 18-base pair palindromes. In this mechanism, the bound EBNA1 then crosslinks EBV DNA to human chromosome 11 [64]. To test this mechanism, existing literature data was first compared to the specific human EBNA1 binding site [65–67]. The results (**Table 4**) are not compatible with a single host sequence that binds EBNA1.

**Table 4.**
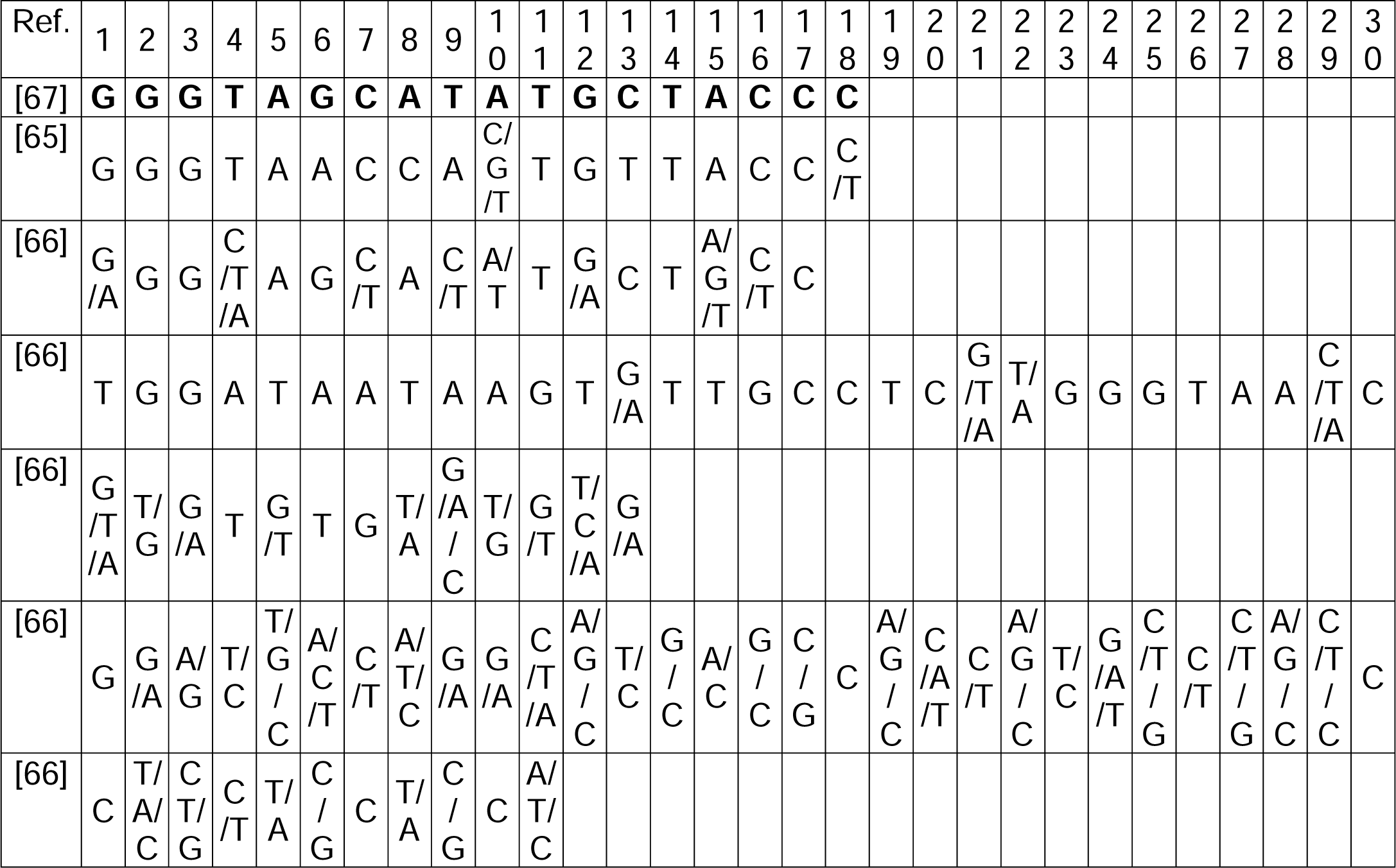
EBNA1-binding sequences in the human genome.

I next measured the probability of finding the imperfect 18 bp palindrome among the 3.2 billion bases of the human genome purely by chance (BLAST analysis "E" (Expect) value). BLAST analysis produced 4,352 matches, from 12 to 18 base pairs in length with E values between 16 and 964 (Supplementary Table S1). Chromosome 11 contained only 197 of these 4352 matches, and none were within or near the palindromic region. The prototype DNA palindrome (**Table 1**, line 2) produced 7074 matches with E values ranging from 0.25 to 964. Further BLAST analyses of the docking sequence in the EBNA1-DNA complex which was crystallized (**Table 4**, line 1) against other genome assemblies [68] revealed matches on chromosomes 2, 19, 4, and 12. Various isolates of human immunodeficiency viruses had 52 matching sequences.

Subsequent experiments tested whether breakpoints at the chromosome 11 palindromic locus occur consistently in EBV-positive lymphoid cancers. I searched for breakpoints in CNVs or SVs in EBV-positive cancers near the palindromic locus (within 10,000 base pairs). The first sequences were from pediatric BL cases. All except one breakpoint were millions of base pairs from the palindromic locus (**Fig. 12**). In a subgroup of 94 EBV-positive BL cases, the palindromic locus was nearly 100 million bp away from the actual major breakpoint coordinates (**Fig. 12B**). Most breakpoints occurred in a narrow region of approximately 36,000 bp (Chr11: 18,915,804-18,951,521). At least 130,000 base pairs still separated the six breakpoints closest to the palindromic hotspot (**Fig 12B**). In 94 BL samples from EBV-positive patients, breakpoints were concentrated within chromosomes 2, 8, 13, 14, and 22 (**Fig. 12C**).

**Fig. 12A.**
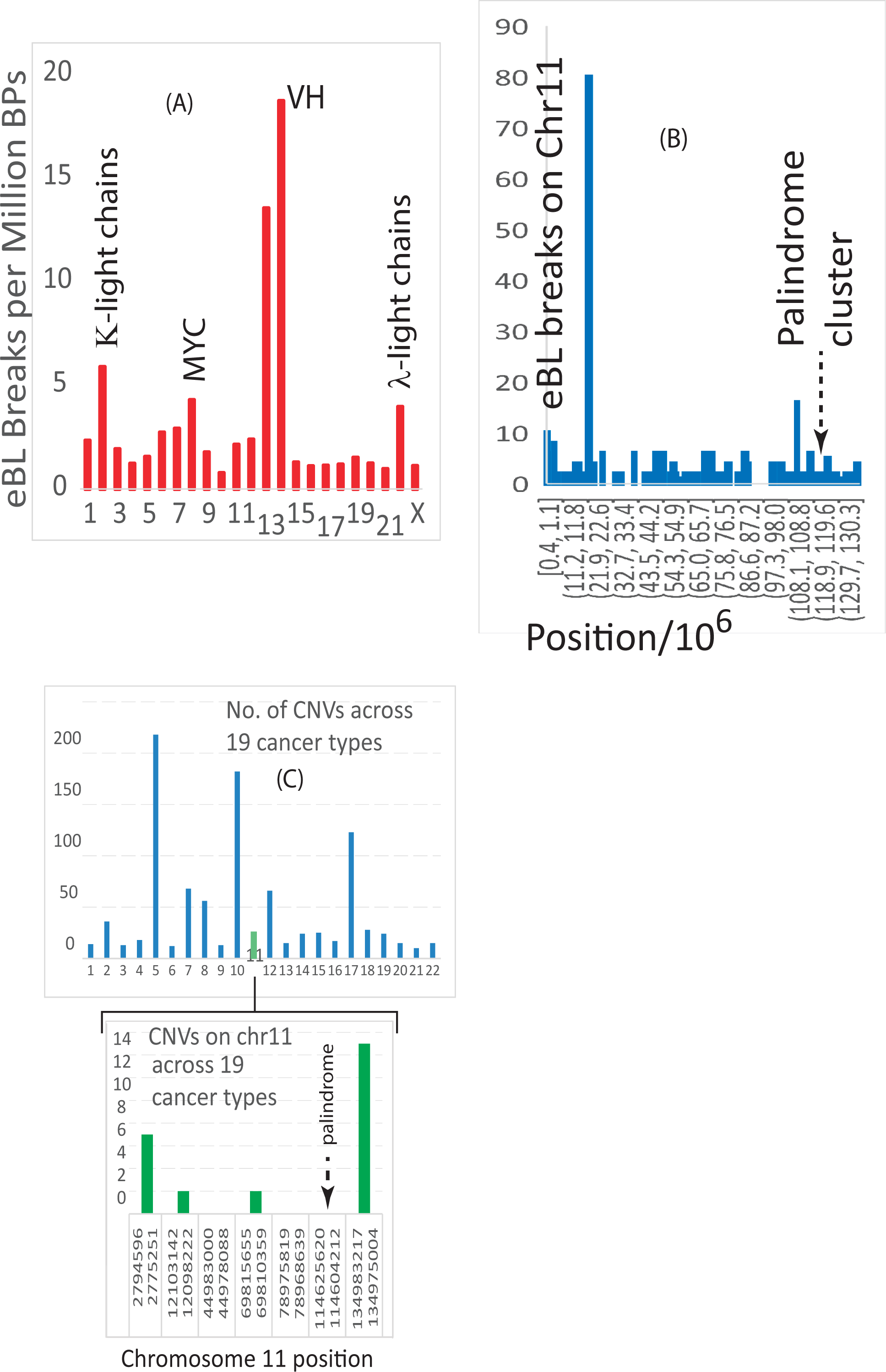
Chromosome breakpoints in 94 patients with EBV-associated endemic Burkitt’s lymphoma (eBL). Fig. 12B. Positions of breakpoints in eBL on chromosome 11. Fig. 12C. Numbers of CNVs on human chromosomes across other cancer types and numbers of CNVs on chromosome 11.

**Fig.13.**
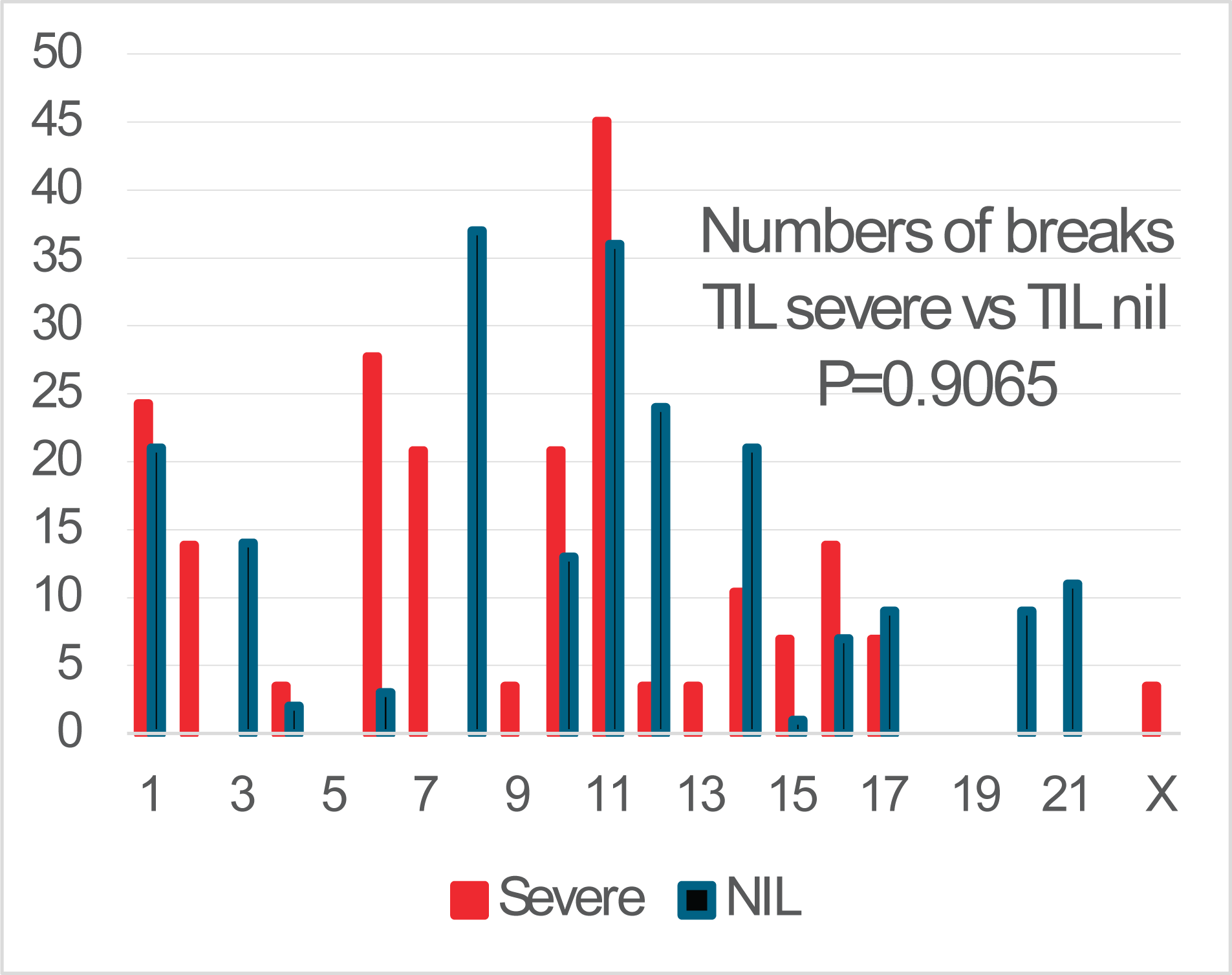
Numbers of breakpoints on chromosomes from breast cancers with severe tumor lymphocyte infiltrates vs. no lymphocyte infiltrates. The normalized values are likely from the same distribution (P=0.9065)

Chromosome 14 contained 610 breakpoints (IgVH regions), and chromosome 2 (IgVK regions) contained 522 breakpoints. EBV hijacks activation induced cytidine deaminase (AICDA) which typically generates antibody gene variants that respond to myriad antigens.

Only 19 of the 94 EBV-positive patients [15] had breakpoints anywhere on chromosome 11 (**Fig 12C**). In BL cells, EBV docks on chromosome 11 near the FAM-D and FAM-B genes [66], but other EBV docking sites on chromosome 11 are over 100 million and 21 million base pairs from the palindrome locus, near the LUZR2 and FAT3 genes, respectively. Therefore, chromosome 11 fusions in the palindromic regions probably do not initiate structural variation in most cases of EBV-positive BL. To test the involvement of the palindromic locus in other cancers, CNVs were compared for diverse cancers from 8227 patients [69]. Neither amplifications nor deletions included the palindromic sequence (**Fig 12C**). The chromosome breakpoints closest to the palindrome locus in 70 NPC cases [16] occur at positions 112,803,697 and 115,316,621.

These breaks are at least 691,000 bp from the palindromic locus. Although the proximity of breast cancer breakpoints to those in NPC on chromosome 11 was clear, the coinciding breakpoints were not near the palindromic locus (**Fig. 6**). All of these results are incompatible with the notion that EBV docking and EBV cancers originate from the crosslinking of EBNA1 and imperfect palindromic repeats on human chromosome 11.

### Tumor-infiltrating lymphocytes do not cause the breaks observed in breast cancer but may influence where they occur

Comparisons of a limited number of breast cancer cases characterized by a large number of tumor-infiltrating lymphocytes (TILs) or no TILs did not find a significant difference in how breakpoints were distributed on the chromosomes (P=0.9073) (**Fig 13**).

### The abundant EBV-like sequences in human chromosomes represent a small section of EBV DNA

To study the feasibility of a future vaccine against EBV, human chromosomes 3, 8, 11, and 17 DNA were arbitrarily selected for comparison to all known and available viral sequences (**Fig. 14**). Many positions along the entire lengths of the four chromosomes resemble EBV and retroviruses, but the distributions of virus-like sequences differ. Chromosome 8 had the strongest resemblance to EBV with 6014 sequences that matched EBV (homology scores over 500, indicating 355-500 identical bases). Because EBV-like sequences are abundant on human chromosomes, EBV can mimic human sequences. The following five EBV-like human sequences have oncogenic properties in NPC [70]:

- LMP2A/LMP2B regulate the host aryl hydrocarbon pathway [71] and maintain latent infection.
- A73 correlates with NPC occurrence [72]
- BALF4 is a virion envelope glycoprotein in spikes that attaches to host cell surface proteoglycans. The viral and cellular membranes then fuse, enabling virus entry. BALF4 dramatically enhances the ability to infect human cells [73]
- BALF3 is an endonuclease that mediates mature virion production and packaging during the EBV lytic cycle[74]. The BALF3 protein causes doublestrand breaks and micronuclei in NPC [75].
- LMP-1 reprograms infected cells, inducing proliferation, inflammation, and preventing differentiation [76]

This result shows that thousands of full or partial copies of EBV-like sequences relate to instability of the human genome (**Fig. 14A** -14C) and emphasizes how dangerous EBV infection can become. Fortunately, only a few areas of EBV genomes were found in human DNA and these sequences were confined to about 2500 bases out of the approximately 173,000 bases in EBV genomes.

**Fig 14A.**
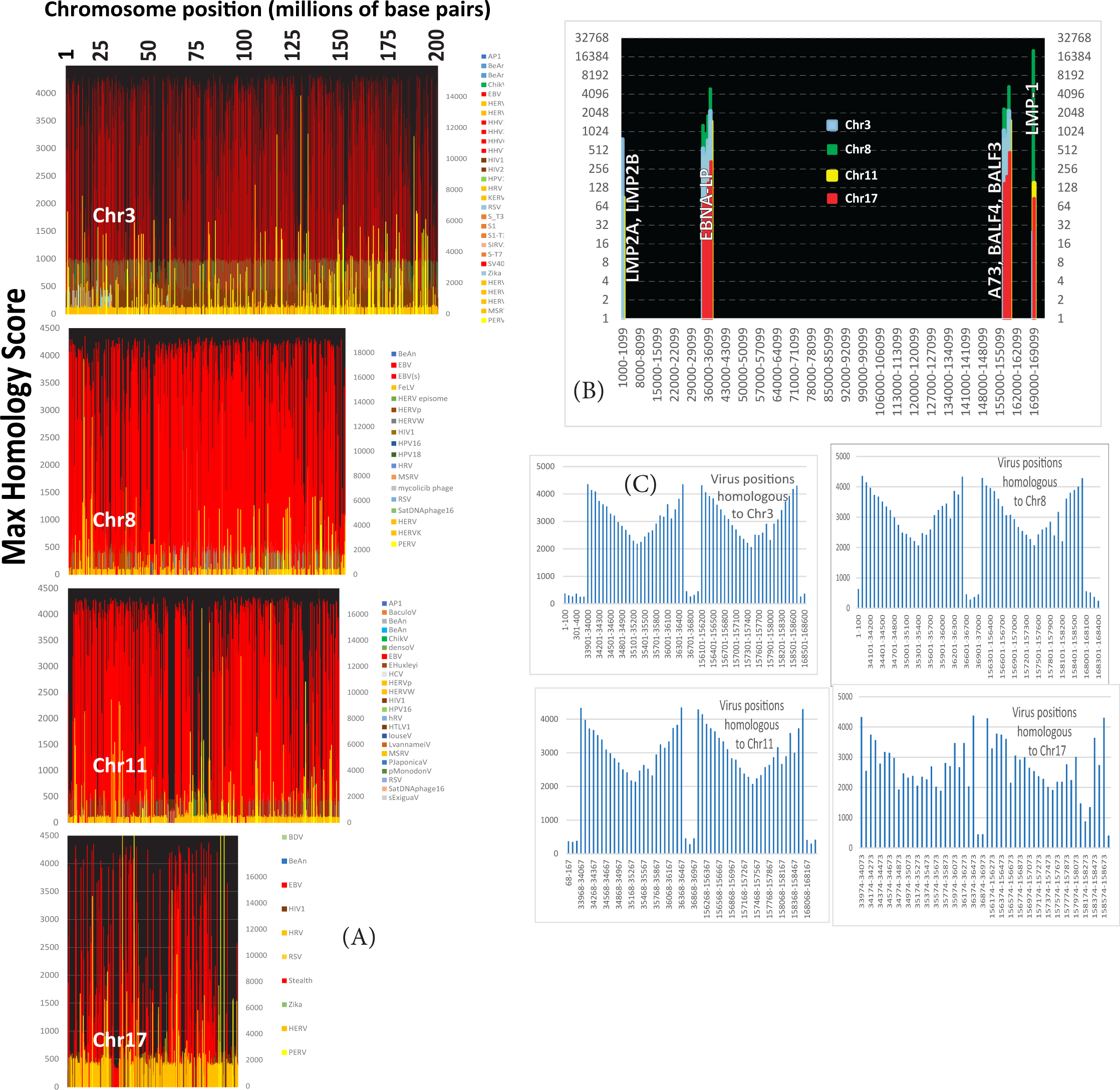
Positions on chromosomes 3, 8, 11, and 17 with significant homology to a known virus. Fig 14B. Genes on the EBV chromosome that correspond to the positions of homology in Fig. 14A. Fig. 14C. Number of homologous sequences at each position in the EBV genome.

## Discussion

This study identified at least five separate lines of evidence that implicate EBV infection in breast cancer (**Fig.15**). First, genome scars left by EBV on breast cancer genomes are characteristic evidence of prior EBV infection, regardless of whether the infection is still active. Breast cancer methylation was observed near the same positions of differential methylation in keratinocytes that had cleared previous EBV infections. This persistent methylation does not occur in never-infected cells.

**Fig. 15.**
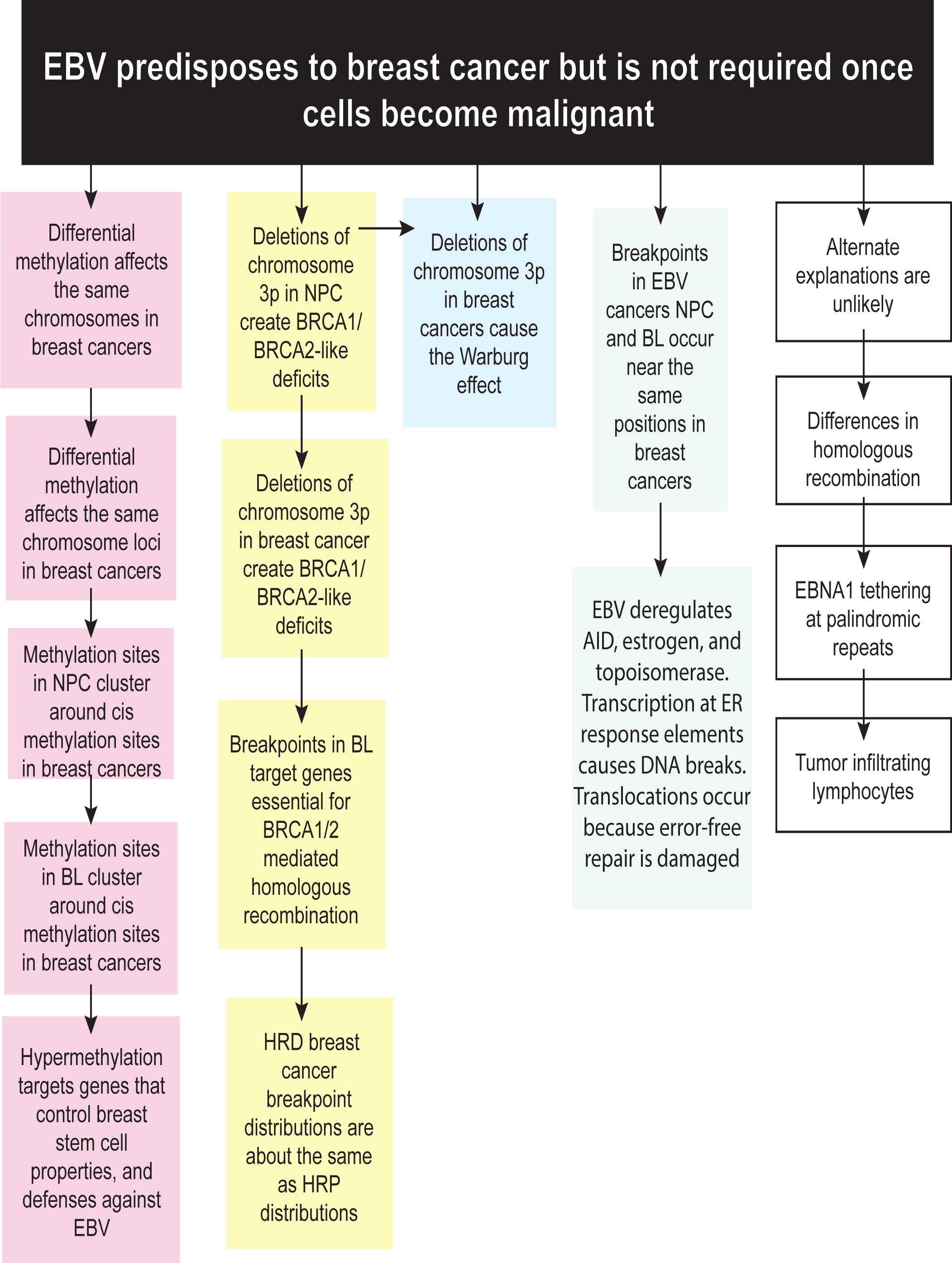
Five independent lines of evidence support the conclusion that EBV predisposes its hosts to breast cancer, but the virus is not required once malignant transformation occurs.

Functional analyses of the genes near these continuing positions suggest that their inhibition could predispose the cells to becoming malignant. Sites of aberrant methylation in NPC and BL, both known EBV-related cancers, occur near sites that are also aberrantly methylated in breast cancers. Stem cell properties, immune responses, and high-fidelity DNA repair are foci of EBV-related methylation that are shared among cancers.

In NPC, near-universal deletions on chromosome 3p remove FANCD2 genes. FANCD2 protein is essential for the function of the breast cancer genes BRCA1 and BRCA2. In NPC, EBV causes deletions, mutations and rearrangements in the host that affect the same HR pathway as hereditary deficiencies in BRCA1 and BRCA2 genes[77–79]. EBV infection in the breast then approximates inheriting a defect in BRCA1 or BRCA2, causing an increased risk for breast cancer. The virus leaves this evidence behind as broken chromosomes, gene rearrangements, deletions, and insertions. The genome positions where these events take place in breast cancer often resemble those in EBV cancers. This result is consistent with the subsequent finding that breakpoints in HRD vs. HRP breast cancers match.

The 3p deletion in EBV cancer also disables normal glucose metabolism via oxidative phosphorylation, causing the Warburg effect. The 3p deletion omits code for PDHB, a key enzyme needed for normal glucose metabolism. Absence of PDHB makes oxidative phosphorylation impossible, so glucose metabolism mimics anerobic conditions in the presence of plentiful oxygen.

Alternate explanations for chromosome breaks and breast cancer invoke initiation by viral tethering at an expandable sequence on chromosome 11. The effect of estrogen is consistent with viral activation of AID because most AID-induced mutations on chromosome 8 in BL are near breast cancer breakpoints. EBV can also deregulate estrogen production, facilitating topoisomerase induced breaks. Finally, abundant EBV-like sequence fragments on at least four chromosomes matched an oncogenic protein from EBV and support the idea that EBV and human chromosomes have extensive interactions.

A strength of this study is its use of alterations to the genome as markers of the presence of a past virus infection. Analysis of data from EBV-infected keratinocytes suggested that EBV could be cleared after multiple generations of breast cancer cell division. The methods and tests developed here should be applied to test other cancers for roles of latent infections. Other cancers also show the Warburg effect and have 3p deletions. For example, small cell lung cancers universally delete part of chromosome 3p [80]. Deletion of chromosome 3p is common in squamous cell cancers [81]. Viral involvement in these cancers should also be tested. A limitation of the current study is the relative rarity of NPC and BL cancers.

The current findings explain inconsistencies in epidemiological studies that evaluated EBV infection in breast cancers. The results of the current study show how EBV can nonetheless contribute to breast cancer even if the infection is ultimately cleared. This contribution becomes independent of the presence of virus because EBV infection leaves marks on the human genome and causes chromosomal aberrations.

The fact that 100% of NPC cases were positive for EBV infection in a selected area of China suggests differences in the role of EBV infection in NPC and breast cancer, because 100% of breast cancer cells do not test positive for EBV infection. These findings may represent geographic differences in NPC prevalence. Geographic differences are also characteristic of breast cancer. Over 2.3 million new cases and 685,000 deaths from breast cancer occurred in 2020. Typical of an infectious disease, the 2.3 million new breast cancer cases reported in 2020 show large variations in different countries and world regions (<40/100,000 females in parts of Asia and Africa to >80 per 100,000 in North America, some areas of Europe, and Australia/New Zealand)[82]. Areas of Asia (prone to NPC) can have over a fifteen fold differences in incidence [83]. However, geographic variation is complex and subject to additional variables such as population density, genetic backgrounds, and environmental factors.

The results presented here predict that vaccination against near-universal EBV infection could markedly reduce the incidence of breast cancer. Breast cancer treatment regimen design may also consider remnants of past EBV infection. However, the significant homology between the human and EBV genomes complicates the development of a vaccine or treatment protocol. The finding that only a few of all possible EBV genetic regions exist in the human genome suggests products of these regions should be excluded from vaccines or treatment protocols. Future research should extend these experiments to the remaining chromosomes.

## Methods

### BL breakpoint data

Breakpoints in endemic BL were obtained from published data from endemic BL (eBL) and sporadic BL (sBL) [15]. Both ends of every CNV and every fusion were counted as breakpoints. Data for comparisons often came from two different genome versions. In these cases, data were converted to GRCh38 coordinates using UCSC LiftOver methods. The length of whole chromosomes was from CHM13 coordinates. Data for 780 breast cancers were obtained from genome sequences compiled from five published studies [54]

### Calculation of distances between breakpoints in different cancers

The break position in a breast cancer genome located closest to a break in an EBV cancer genome (NPC or BL) was taken as the XLOOKUP value for the number of base pairs from the closest NPC breakpoint 5’ or 3’ to the BL break or the NPC breakpoint (Supplementary Table S2). The distance from the breast cancer breakpoint was then the absolute value of the difference between the closest EBV cancer break and the tested break in breast cancer.

Differences in the amount of data available for NPC, BL, and breast cancers complicated the calculations, especially near telomeres. Several methods of handling these end regions were tried and made no meaningful difference in the outcomes.

Several different window sizes were tried for making comparisons of positions in breast cancer vs. EBV-related cancers. A 10,000 base pair window with an overflow window of 10,000,000 was used for initial screening, and often proved to be adequate.

Later, an optimum window size used was calculated from a least squares plot of distances of base pairs vs numbers of breakpoints. The window size was taken as the last point that gave a consistent slope. These results gave a value of approximately 5000 for both chromosome 1 and chromosome 11, so 5000 was used as the default window size. For these two chromosomes, the ratio of chromosome breaks in breast cancers nearest NPC to the ratio of chromosome breaks nearest randomly generated values were maximum near a window size of 5,000. In contrast, translocation breakpoints on chromosome 8 comparing MYC and BL were linear up to a value of 10,000 bp and then leveled off. Alternative methods of calculation using array formulas always gave the same results as the XLOOKUP methods. The minimum distances separating breast cancer breaks from EBV cancer breaks in these arrays were calculated as the absolute value of breast cancer break position minus the array of EBV cancer break positions. The millions of calculations required were repeated at least twice and often many times (supplementary Table 2)

### Comparisons of distributions

Tests for normality routinely showed that the data was unlikely to be normally distributed. Therefore, Mann-Whitney nonparametric tests were regularly used to compare the distributions of breakpoints and methylated promoters. Correlation between epigenetic modifications on breast vs EBV cancers was by linear regression analyses with a power for 5% significance of 99.99%. Kolmogorov-Smirnov and Spearman’s tests were also used. Frequencies were plotted to verify the distributions of coinciding areas.

### DNA sequence homology

The NCBI BLASTn program (MegaBLAST) and database [84–86] were used to compare the consensus DNA palindrome sequence to human DNA sequences and to determine the homology of human DNA to viral DNA. BLAST corrects for short input sequences by using a smaller word size and a different scoring algorithm. E(expect) values are related to p values and represent the probability that a given homology bit score occurs by chance. A strict cutoff was used because the 3.2 billion bases in the human genome increase the odds that identical sequences can occur by chance. Only E values <1e-10 were considered to indicate significant homology. In cases with significant homology, the E values were represented as "0" (<1e-180) and were always far below 1e-10.

## Supporting information

supplementary table 1 EBNA1 homologies

supplementary Table 2 sample calculations

## Data Availability

All data produced in the present study are available upon reasonable request to the authors

---

**Supplementary Fig. 1.**
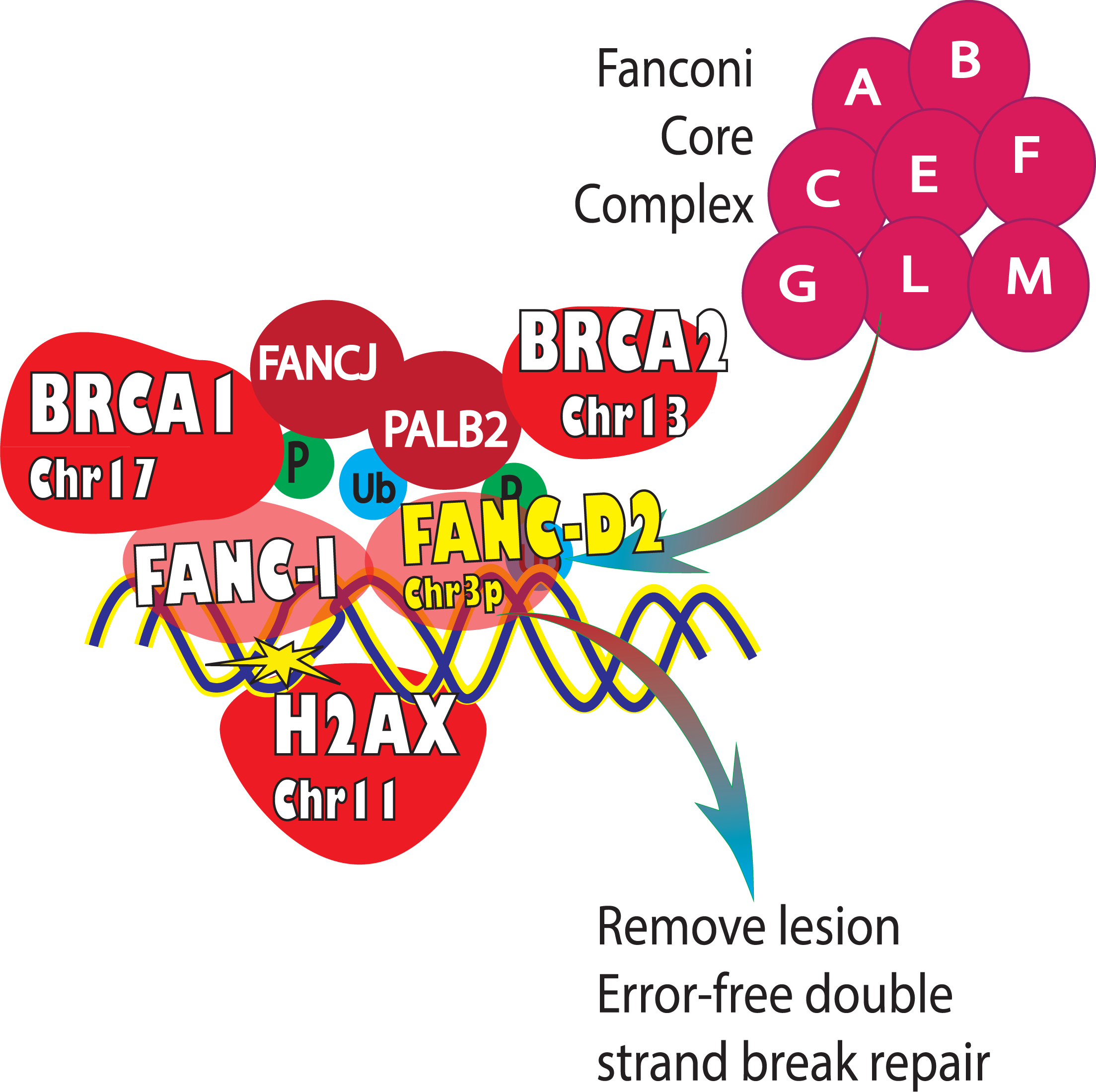
Summary of selected gene product interactions in the pathway for error-free double strand break repair. FANCD2 is a critical intermediate encoded on chromosome 3p. The pathway is based on many publications of D’andrea et al. (e.g. references 30, 62)

